# Complexome profiling showed impaired immunoproteasome assembly in a novel PRAAS subtype caused by monoallelic PSMB8 variants

**DOI:** 10.1101/2025.06.25.25330124

**Authors:** Robin Wijngaard, Caspar I. van der Made, Sema Kalkan Uçar, Gayatri Ramakrishnan, Jill A. Rosenfeld, Tiphanie P. Vogel, Sarah K. Nicholas, Monika Weisz-Hubshman, Undiagnosed Diseases Network, Alfredo Cabrera-Orefice, Bente Siebels, Thomas Mair, Hartmut Schlüter, Ruben L. Smeets, Ronald van Beek, Clara D.M. van Karnebeek, Alexander Hoischen, Lisenka E.L.M. Vissers, Saskia B. Wortmann, Ron A. Wevers, Machteld M. Oud, Sergio Guerrero-Castillo

## Abstract

Immunoproteasomes, essential for MHC class I antigen presentation, differ from standard proteasomes by incorporating the catalytic subunits PSMB9 (β1i), PSMB10 (β2i), and PSMB8 (β5i). Proteasome-associated autoinflammatory syndromes (PRAAS) are type I interferonopathies resulting from impaired proteasome function. Here, we describe two individuals carrying monoallelic *de novo* variants in *PSMB8*, both presenting with early-onset systemic autoinflammation and features of immunodeficiency, accompanied by a marked type I interferon response. To investigate the underlying mechanism, we performed complexome profiling on interferon-γ-stimulated fibroblasts harboring the p.(Ala235Asp) variant. This variant, located at the β-ring interface, disrupted proper assembly of the immunoproteasome, resulting in reduced levels of fully assembled 20S and 26S immunoproteasomes and accumulation of assembly intermediates. The findings suggest a dominant-negative effect and broaden the clinical and genetic spectrum of PRAAS with immunodeficiency (PRAAS-ID), while highlighting the utility of complexome profiling to study proteasome assembly defects.

## Introduction

Proteasomes are large multiprotein complexes that mediate protein degradation within cells. They primarily target ubiquitinated proteins but also process non-ubiquitinated substrates (1). The proteasome is a dynamic protein complex composed of a catalytic 20S core particle combined with up to two additional subcomplexes, one at each end, regulating its activity (1–3). The 20S core particle is a cylindrical structure of 28 subunits, which serves as the central proteolytic unit. It is composed of four stacked rings: two outer rings of seven α-subunits and two inner rings of seven β-subunits (4). The inner β-rings of the standard proteasome (SP) contain non-catalytic as well as three catalytic subunits, PSMB6 (β1), PSMB7 (β2), and PSMB5 (β5), which exhibit caspase-like, trypsin-like, and chymotrypsin-like activity, respectively (5). The catalytic subunits are synthesized as precursors and activated through N-terminal propeptide cleavage, exposing the catalytic threonine residues at the active sites. In addition to the constitutively expressed SP, tissue-specific forms exist, including the immunoproteasome (IP), the thymoproteasome and the spermatoproteasome. The IP is expressed in immune cells but can also be induced in other cell types in response to interferon-γ (IFNγ) or other pro-inflammatory cytokines (6, 7). The IP has distinct peptide cleavage properties that are optimized for generating antigenic peptides for MHC class I binding, contributing to the adaptive immune response (8–10). In the IP, the three catalytic subunits found in the SP are replaced by the specialized subunits PSMB9 (β1i), PSMB10 (β2i), and PSMB8 (β5i) (11).

The assembly of the 20S proteasome begins with the formation of the α-ring, and, subsequently, the β-subunits are stepwise incorporated (2, 12, 13). The structure of one α-ring and one β-ring is known as the 13S intermediate or half-mer. This half-mer is dimerized with a second half-mer to form the complete 20S proteasome. The assembly process is tightly regulated and requires multiple dedicated factors, such as assembly chaperones PSMG1-4 or POMP (12). These chaperones disassociate from the proteasome as assembly progresses and are absent in the mature complexes (12). Although the assembly mechanism is largely conserved, some differences have been reported between the assembly of the 20S of the SP and the IP (2, 6). In the IP, PSMB9 is incorporated earlier in the process than its homologous subunit, PSMB6, in the SP. This leads to the formation of an incomplete intermediate containing one α-ring and the PSMB9, PSMB10, PSMB3 (β3) and PSMB2 (β4) subunits, preceding the incorporation of the remaining subunits, including PSMB8 (13, 14). The 20S proteasome of both the SP and the IP can bind to one or two identical or distinct regulatory particles, including the 19S (PA700), the 11S (PA28), or the PA200 regulatory particle (1).

Dysfunction of the IP gives rise to a group of inborn errors of immunity known as proteasome-associated autoinflammatory syndromes (PRAAS). PRAAS are type I interferonopathies due to sustained pathological expression of type I interferon cytokines (15, 16). Chronic activation of the interferon pathway leads to an increased expression of interferon-stimulated genes, referred to as the type I interferon signature, which can be measured and serves as a diagnostic biomarker (16). The clinical phenotype consists of multisystemic hyperinflammation, including symptoms such as rash, fevers, and organ dysfunction. Most PRAAS disorders are inherited in an autosomal recessive manner or follow oligogenic models involving two or more variants in different subunits (17–21), affecting IP catalytic subunits, assembly chaperones or shared proteasome subunits (MIM #256040, #617591, #619183, #619175). In contrast, autosomal dominant inheritance has been described for variants in the 20S chaperone *POMP* (MIM #618048), and in the IP-specific subunits *PSMB9* (MIM #620796) and *PSMB10* (MIM #620807) (22–24). In these cases, the variants are thought to act via a dominant-negative mechanism, interfering with the normal proteasome assembly.

In this study, we describe two individuals harboring *de novo* variants in *PSMB8*, resulting in a PRAAS-like disorder characterized by systemic hyperinflammation with immunodeficiency. Using complexome profiling, we investigated the functional impact of one of these variants, revealing that it disrupts the normal assembly of the 20S core complex, specifically affecting the IP. These findings support the hypothesis that monoallelic *PSMB8* variants cause PRAAS with immunodeficiency through a dominant-negative mechanism and highlight the utility of complexome profiling in studying proteasome assembly and dysfunction.

## Results

### *De novo* variants in *PSMB8* lead to a PRAAS-like syndrome with immunodeficiency

We identified two unrelated individuals presenting with sporadic early-onset multisystemic hyperinflammation accompanied by immunodeficiency (Tables 1 and 2 and Supplementary Table 1). Both individuals first presented with an infection and multi-organ inflammation including elevated inflammatory markers, liver dysfunction with transaminitis, enteropathy, and myositis with transient creatine kinase (CK) elevations. Both individuals had prominent lung disease, necessitating lung transplantation in individual 2 due to severe bronchitis and bronchi(ol)ectasis. In individual 1 additional cardiac involvement with pulmonary hypertension and mild tricuspid regurgitation was seen. Individual 1 experienced periodic exacerbations of disease symptoms, manifesting as recurrent infections, fevers, and rash with erythematous and edematous plaques. Individual 2 additionally showed failure to thrive, growth hormone deficiency and had a mild neurocognitive disorder, bilateral conductive hearing loss and foci of T2 hyperintensity in the subcortical and white matter (without intracerebral calcification by brain MRI).

**Table 1.**
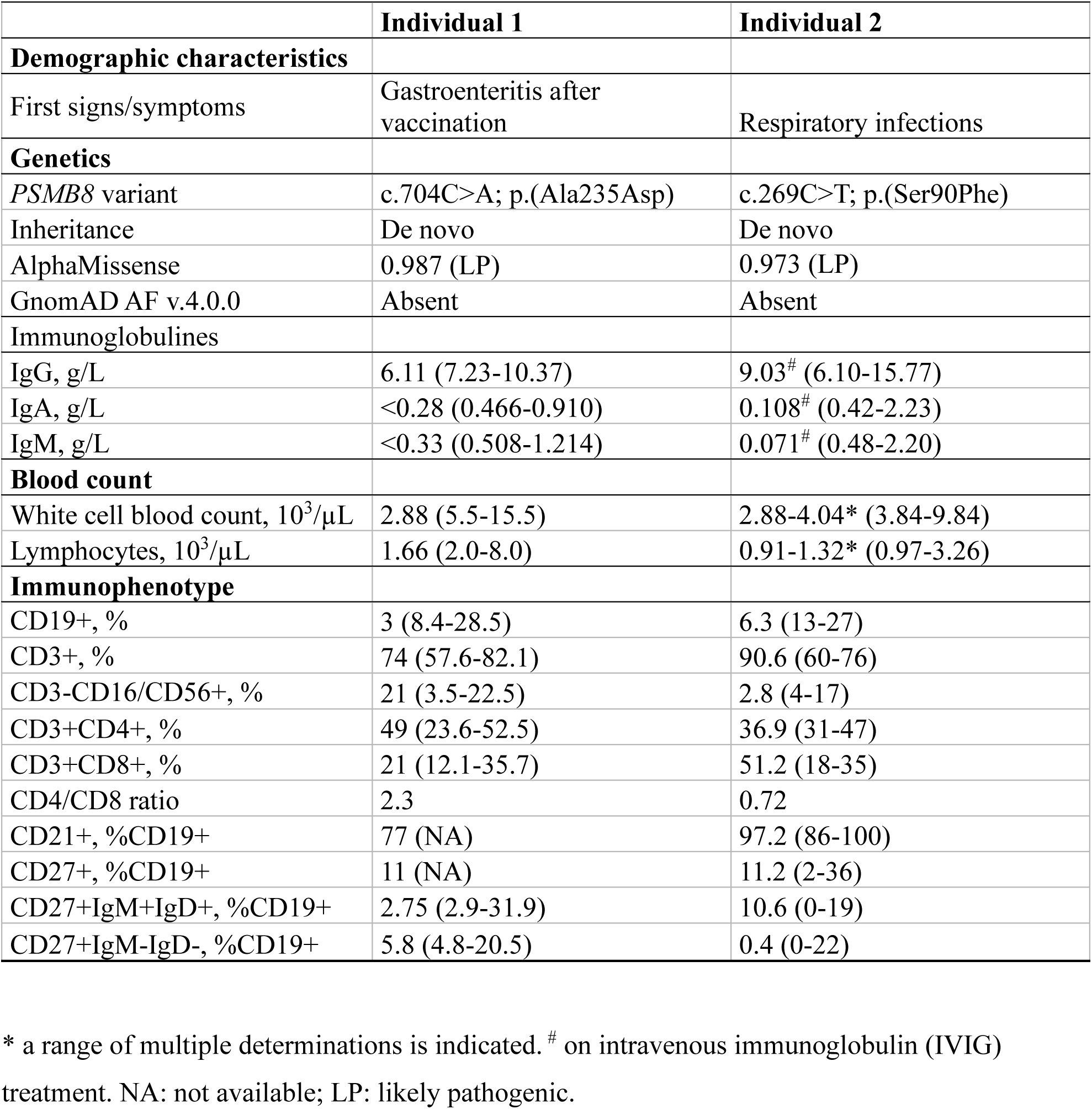
Genetics and immunological laboratory findings of included individuals.

**Table 2.** Comparison of clinical features between monoallelic PSMB8 disorder and other PRAAS syndromes.

| **Disease** | **Monoallelic** |  |  |  |  | **Biallelic, oligogenic** |  |
| --- | --- | --- | --- | --- | --- | --- | --- |
| **Gene(s)** | **PSMB8**<br>**(individual 1)** | **PSMB8**<br>**(individual 2)** | **PSMB9** | **PSMB10** | **POMP** | **PSMB8** | **PSMB8 (oligo),**<br>**PSMB10,**<br>**PSMB4,**<br>**PSMG2,**<br>**PSMA3,**<br>**PSMA5,**<br>**PSMC5** |
| References | This study | This study | (23, 35) | (24) | (20, 22, 59) | (21, 60, 61) | (18, 20, 21, 62) |
| Total individuals | 1 | 1 | 3 | 6 | 4 | 20 | 13 |
| **Clinical Features** |  |  |  |  |  |  |  |
| Growth deficiency | - | + | NR | + | +/- | + | + |
| Recurrent fever | + | - | + | - | +/- | + | + |
| Rash | + | - | + | + | + | + | + |
| Systemic inflammation | + | - | + | +/- | +/- | + | + |
| Inflammatory lung disease | + | + | + | +/- | +/- | +/- | +/- |
| Pulmonary hypertension | + | - | + | - | NR | NR | NR |
| Enteropathy | + | + | NR | + | +/- | NR | NR |
| Liver dysfunction | + | + | + | + | - | +/- | + |
| Myositis/Muscle atrophy | + | + | + | - | - | +/- | + |
| Rhabdomyolysis | + | - | + | - | - | +/- | +/- |
| Basal ganglia calcification | - | - | + | - | NR | +/- | +/- |
| Lipodystrophy | - | - | - | - | +/- | + | + |
| Joint contractures | - | - | NR | - | - | +/- | + |
| **Laboratory results** |  |  |  |  |  |  |  |
| Dyslipidemia | + | + | - | NR | NR | + | + |
| Autoantibodies | Neg | NR | Neg | NR | Pos | Pos (mostly) | Pos (mostly) |
| Immunoglobulins | ↓ | ↓ | ↓ | ↓ | ↑ | ↑ (mostly) | ↑ (mostly), ↓ (some) |
| Anemia | + | + | + | NR | + | + | + |
| Thrombocytopenia | + | + | + | NR | + | +, some ↑ | +, some ↑ |
| B cells | ↓ | ↓ | ↓ or normal | ↓ | ↓ | Normal or NR | Normal or NR |
| T cells | Normal | Normal | ↓ or normal | ↓, ↑ CD4/CD8 | ↑ CD4,<br>↑CD4/CD8 | Normal or NR | Normal or NR |
| NK cells | Variable | Variable | Variable | Variable | ↓ | Normal or NR | Normal or NR |
| IFN signature | ↑ | ↑ | ↑ | ↑ | ↑ | ↑ | ↑ |
| Other |  |  | Abnormal coagulation | Eosinophilia | Eosinophilia | Low TSH (uncommon) |  |

-: not observed; +/-: observed (some patients); +: observed (most patients), NR: not reported; Pos: positive; Neg: negative

Microcytic anemia, (intermittent) thrombocytopenia, and lymphopenia were observed in both individuals. Immunophenotyping revealed significant and repeated reductions in B cell percentages, particularly affecting the proportion of switched memory B cells (%CD19+CD27+IgM-IgD-) in individual 2 (Table 1). Individual also 2 presented low circulating NK cell numbers. In contrast, T cells were normal in both number and function, with normal responses observed after stimulation with phytohemagglutinin (PHA). Additional immunological analyses demonstrated hypogammaglobulinemia, necessitating ongoing intravenous immunoglobulin (IVIG) therapy in both individuals. Interferon scores were markedly elevated in individual 1 and mildly elevated in individual 2, although this measurement was performed during immunosuppressive treatment following lung transplantation (Supplementary Figure 1). In line with the elevated interferon signature, evaluation of soluble serum factors revealed substantial elevations (>1.5× ULN) in individual 1 for the following markers: BCA-1, MIP-3, MIP-4, MIG, IFN-γ, IL-18, IP-10, IL-6, IL-8, sVCAM-1, sTNFR2, and sIL-2Rα (Supplementary Table 2). These findings suggest marked activation of the T-lymphocyte compartment (e.g., sIL-2Rα), particularly the Th1–IFN-γ pathway (e.g., IL-18, IFN-γ, IP-10, MIG), along with broader inflammatory activity, as indicated by the other markers. Autoantibody panels were negative in Individual 1 (Supplementary Table 3). A combination of clinical features and laboratory findings including fever, bicytopenia, hyperferritinemia, splenomegaly and hypertriglyceridemia in this individual raised a suspicion of hemophagocytic lymphohistiocytosis (HLH).

### Genetic analysis identified a *de novo* variant in *PSMB8*

The genetic variants of the individuals were analyzed and assessed by their respective centers. For both individuals, a missense variant was identified in *PSMB8:* NM_148919.4:c.704C>A; p.(Ala235Asp) for individual 1 and NM_148919.4:c.269C>T; p.(Ser90Phe) for individual 2. The variant was confirmed to be *de novo* in both cases (Supplementary Figure 2A-B). Both variants were absent from gnomAD v4.0.0 and were predicted to be deleterious by multiple *in silico* predictors including AlphaMissense (Table 1). Given the potential for digenic inheritance, all other subunits of the proteasome complex were screened for variants. No other rare exonic variants were identified in individual 1 nor 2, but only inherited intronic or upstream variants (Supplementary Table 4). SpliceAI predicted that these variants would not affect splicing. Moreover, reverse-transcriptase PCR of RNA isolated from fibroblasts of individual 1 and RNA-seq of individual 2 confirmed that these intronic variants did not affect splicing, suggesting that they were unlikely to contribute to the observed phenotype (data not shown).

### The identified *PSMB8* variants destabilize the 20S and 26S complexes *in silico*

We investigated the structural impact of variants p.(Ala235Asp) and p.(Ser90Phe) in the PSMB8 subunit of the 20S and 26S IP complexes through *in silico* structural modeling. Both affected residues in PSMB8 are retained in the mature protein and are conserved across species (Figure 1A).

**Figure 1.**
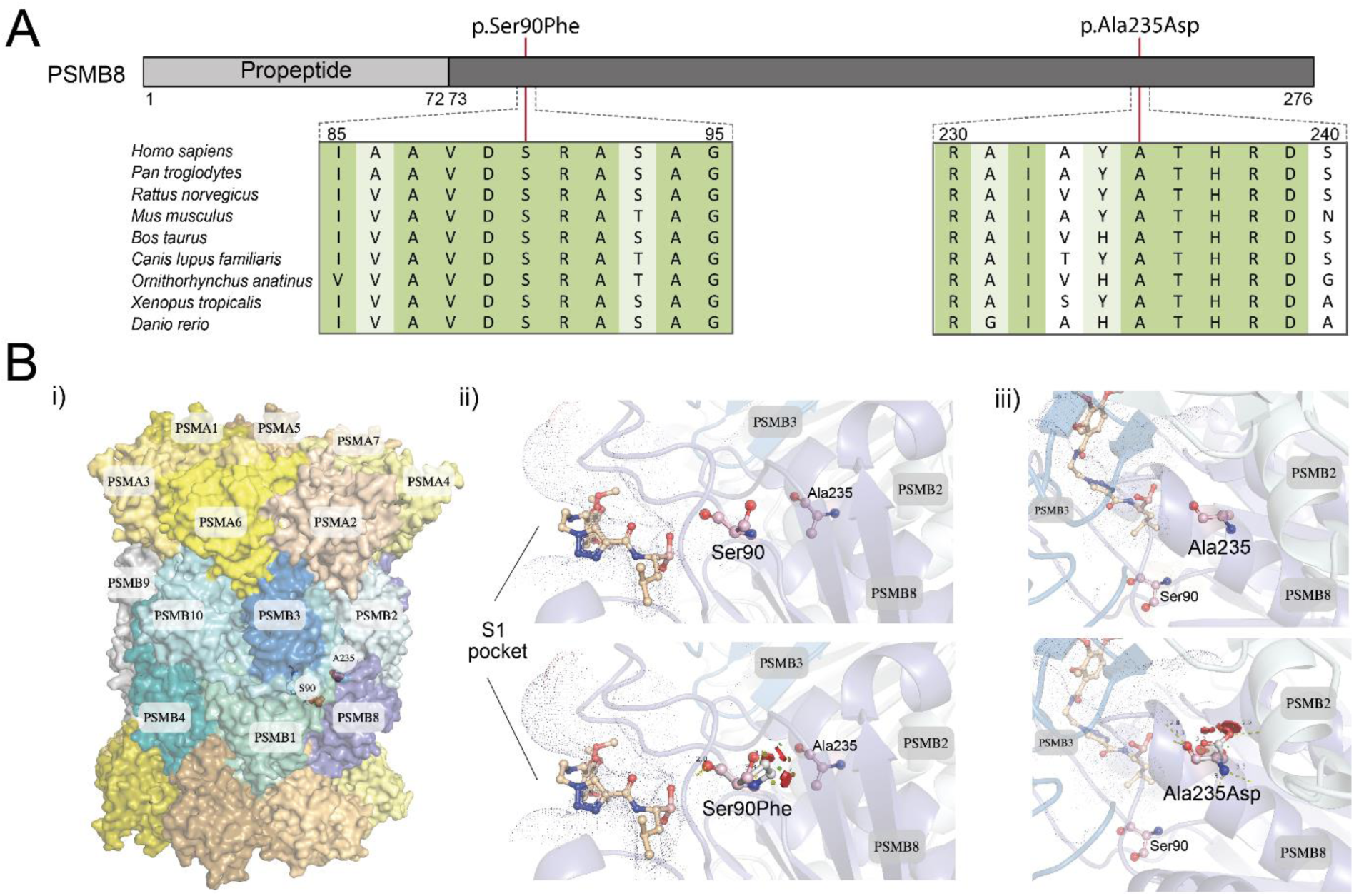
Structural modeling of the variants. **A:** Multiple sequence alignment of the PSMB8 subunit across species. Highly conserved residues are shown in green, and moderately conserved residues in light green. **B**: Structural modelling of the two variants. i) Crystal structure of the 20S immunoproteasome assembly with the variants indicated. ii) Close-up view of the p.(Ser90Phe) variant located near the S1 pocket of the active site. iii) Close-up view of the p.(Ala235Asp) variant located at the PSMB2-PSMB8 interface. Both variants introduce steric clashes in the neighborhood indicated as red discs.

Ala235 is located near the PSMB8–PSMB2 interface formed between the two opposing β-rings of the 20S proteasome, at a conserved hydrophobic site thought to be structurally important. The p.(Ala235Asp) substitution introduces steric clashes within the complex, likely disrupting subunit interactions (Figure 1B). Moreover, the negatively charged Asp residue is incompatible with the hydrophobic environment. The substitution is thus predicted to be highly destabilizing within the 20S structure (ΔΔG_mut-wt_ = 28.26 kcal/mol) and the 26S proteasome structure (ΔΔG_mut-wt_ = 16.91 kcal/mol).

In contrast, Ser90 does not interact with neighboring subunits but is located near the S1 pocket of the active site (Figure 1B) and the propeptide cleavage site (25). Evolutionarily, this position was identified to be conserved across species (Figure 1A) and can only accommodate Ser or Thr, suggesting a functional role within the protein. Moreover, Ser90 might have allosteric interactions with residues lining the S1 pocket. The substitution to a bulky and hydrophobic Phe likely imparts rigidity to the active site loop, involved in the protein function. Similar to p.(Ala235Asp), the p.(Ser90Phe) variant also exhibits destabilizing effects in both the 20S (ΔΔG_mut-wt_ = 13.05 kcal/mol) and 26S complexes (ΔΔG_mut-wt_ = 6.13 kcal/mol), indicating potential deleterious impacts on structural integrity.

Both the variants showed substantial shifts in local energy frustrations. Residues in contact with p.(Ala235Asp), within a sphere of 5Å centered at Cα of Ala235, shifted from a neutral/minimally frustrated state to a highly frustrated state. A total of 10 out of 56 pairs of contacts, involving 5 of 12 interacting residues (41.67%), are predicted to undergo this shift (Supplementary Figure 3B, Supplementary Table 5). Notably, most of 56 contact pairs were in either a neutral or minimally frustrated state, thereby making this shift potentially unstable. In contrast, Ser90Phe introduced shifts from a neutral/highly frustrated state to minimally frustrated state affecting 16 out of 44 contact pairs, involving 9 of 36 interacting residues (25%) (Supplementary Figure 3C, Supplementary Table 5). In this case, we identified an increase in the number of minimally frustrated contacts, consistent with the FoldX estimate on p.(Ser90Phe) imparting rigidity to the loop, and potentially altering local structure and function.

The recessive variants p.(Ala92Thr), p.(Ala92Val), p.(Ala94Pro), p.(Met117Val), and p.(Ser118Pro) were analyzed for comparison, as they are clustered near the active site in close proximity to Ser90 (Supplementary Figure 3A). Contrary to the dominant variants, these substitutions caused only marginal changes in local frustration, affecting few residues in the neighborhood. Interestingly, this differs from the FoldX results, which predict p.(Met117Val) and p.(Ser118Pro) to be destabilizing variants (Supplementary Table 5). This discrepancy suggests that these recessive variants may induce localized effects at the active site, rendering the protein non-functional while largely preserving the overall structural integrity of the complex.

### Complexome profiling enables analysis and differentiation of IP from SP

To determine whether IP 20S and 26S complexes were properly assembled in fibroblasts from individuals harboring a variant in *PSMB8*, complexome profiling was employed. This mass spectrometric technique separates protein complexes under native conditions to analyze their composition and abundance within a biological sample.

The ability of this technique to detect all proteasome subunits and to differentiate between the SP and the IP was first assessed using a non-immune cell line (143B) and an immune cell line (THP-1). The migration profiles in these cell lines showed two predominant peaks at ∼750 kDa and ∼2000 kDa, corresponding to the 20S and 26S complexes, respectively (Supplementary Figure 4A). At the 20S peak, seven α and ten β-subunits were detected, including PSMB5-7 (SP-specific) and PSMB8-10 (IP-specific), indicating the presence of both proteasome types in immune and non-immune cell lines. The 26S complex additionally included ATPase and non-ATPase regulatory subunits from the 19S regulatory particle. A peak around ∼1000 kDa, close to the 20S proteasome, was also detected, representing the 20S complex associated with the 11S or PA200 regulatory subunits. Additional peaks corresponding to proteasome subcomplexes were observed and are further detailed in the Supplementary Results and Supplementary Data 1.

To differentiate the SP from the IP, we used abundances of PSMB5-7 and PSMB8-10 as SP-specific and IP-specific markers, respectively (Supplementary Figure 4B). In the 143B cells, 95% of the proteasome corresponded to the SP, whereas in the THP-1 cells, the ratio was inverted, with 80% of the proteasome corresponding to the IP (Supplementary Figure 4C). Based on these findings, although the apparent molecular masses of the 20S and 26S complexes of SP and IP were similar, we were able to distinguish between these two based on the quantification of the respective specific β-subunits.

### IFNγ stimulation enhances IP expression in fibroblasts

To better characterize the assembly of the IP in non-immune cell lines, such as fibroblasts, we enhanced the expression of IP-specific subunits via IFNγ stimulation. In untreated control fibroblasts, the abundance of SP-specific subunits PSMB5-7 was higher than the IP-specific subunits PSMB8-10, which only represented 15% (Supplementary Figure 5A-C). Stimulation of these fibroblasts with IFNγ led to an increased abundance of PSMB8-10 to levels compared to PSMB5-7, raising the proportion of IP to 65% (Supplementary Figure 5A-C).

### The IP-specific 20S and 26S complexes are less abundant in fibroblasts harboring p.(Ala235Asp)

Fibroblasts from individual 1 harboring the p.(Ala235Asp) variant in PSMB8 were analyzed by complexome profiling after IFNγ stimulation to investigate whether IP assembly was impaired. The heatmap of α- and β-subunits revealed distinct peaks corresponding to the 20S proteasome, the 20S–11S and/or 20S–PA200 complexes, and the 26S proteasome at their expected molecular weights (Figure 2A). Notably, the abundance of both 20S proteasomes, including those associated with 11S and/or PA200, and 26S proteasomes was reduced in the p.(Ala235Asp) cell line compared to fibroblasts from two healthy controls (Figure 2C–D).

**Figure 2.**
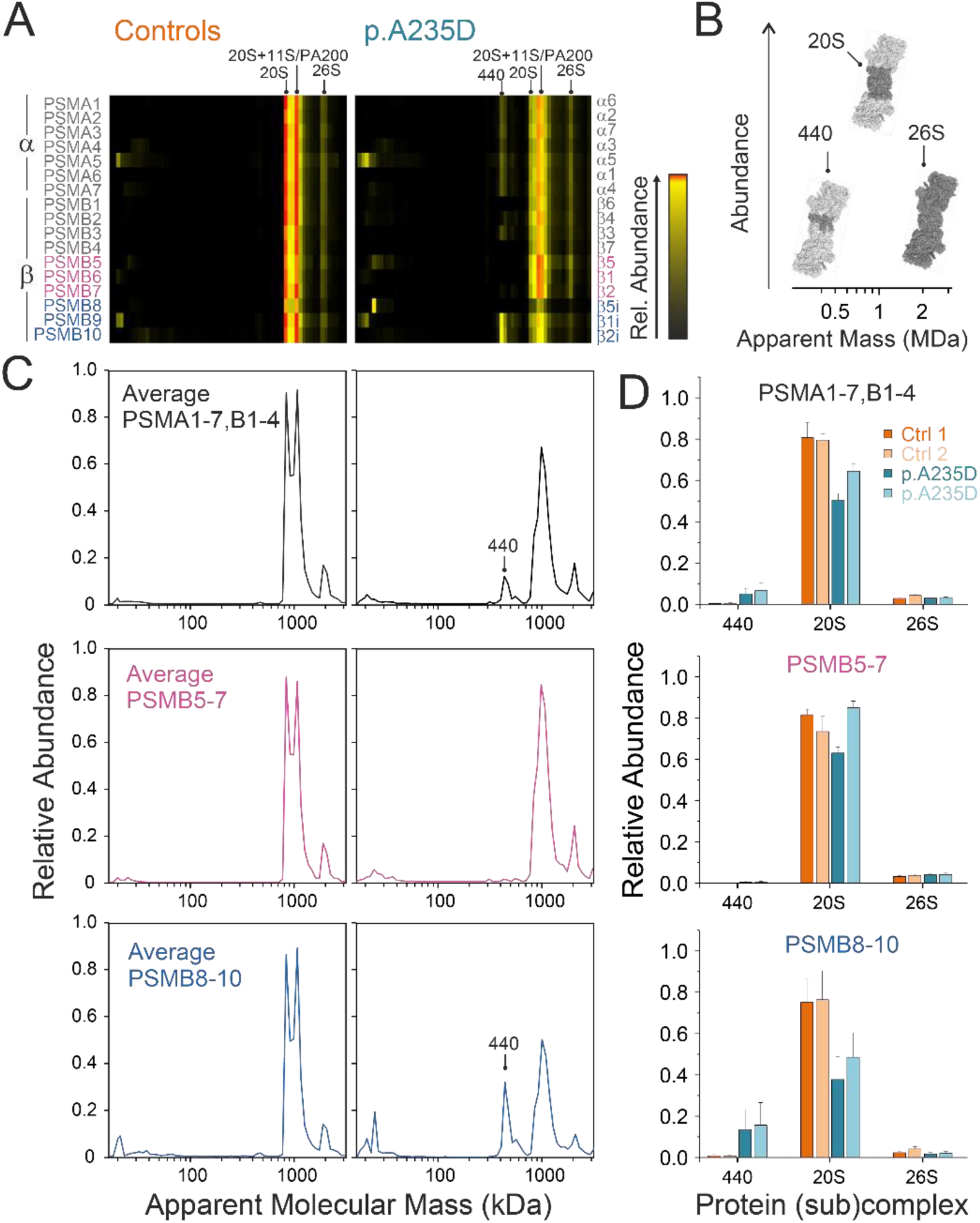
Impaired assembly of immunoproteasomes in the p.Ala235Asp cell line of individual 1. **A**: Heatmap representation of the migration profiles of α- and β-subunits showing signals for 20S* and 26S proteasomes in the control and p.(Ala235Asp) cell lines and an additional signal at ∼440 kDa in the p.(Ala235Asp) cell line. Average of two independent experiments. **B**: Cartoon representation of the complexes identified by complexome profiling. Protein components of each complex are highlighted in dark gray on the cryo-EM structure of the human 26S proteasome, PDB: 5JGR (63). **C:** From top to bottom: average migration profiles of α and β subunit shared between the SP and IP; SP-specific β subunits; and IP-specific β subunits. **D:** Quantification of α- and β-subunits grouped according to panel C at the mass ranges of the ∼440 kDa subcomplex, the 20S and the 26S proteasomes (Mean ± standard deviation, n=2). Mass ranges used for quantification were as follows: 440 kDa peak: 403-514 kDa; 20S proteasome: 774-1262 kDa; 26S proteasome: 1898-2059 kDa. *Note: Quantification of the 20S proteasome also includes 20S incorporated into 20S–11S and/or 20S–PA200 assemblies, as these complexes were not fully separated in the cell lines.

Upon further analysis, we found that only PSMB8-10 showed a 50% reduction of abundance at the 20S and 26S peaks compared to controls while PSMB5-7 showed no change. This suggests that only the IP was affected, while the abundance of the SP remained comparable between individual 1 and the controls (Figure 2D). The decrease of the 20S complex in individual 1 was also observed by comparing the abundance of each subunit separately (Figure 3A-B). Furthermore, in the case of the 26S complex, although no significant difference was observed, the abundance of all shared and IP-specific subunits seemed to be lower in individual 1. No differences were detected in the subassemblies of the regulatory particles (Supplementary Figure 6A).

**Figure 3.**
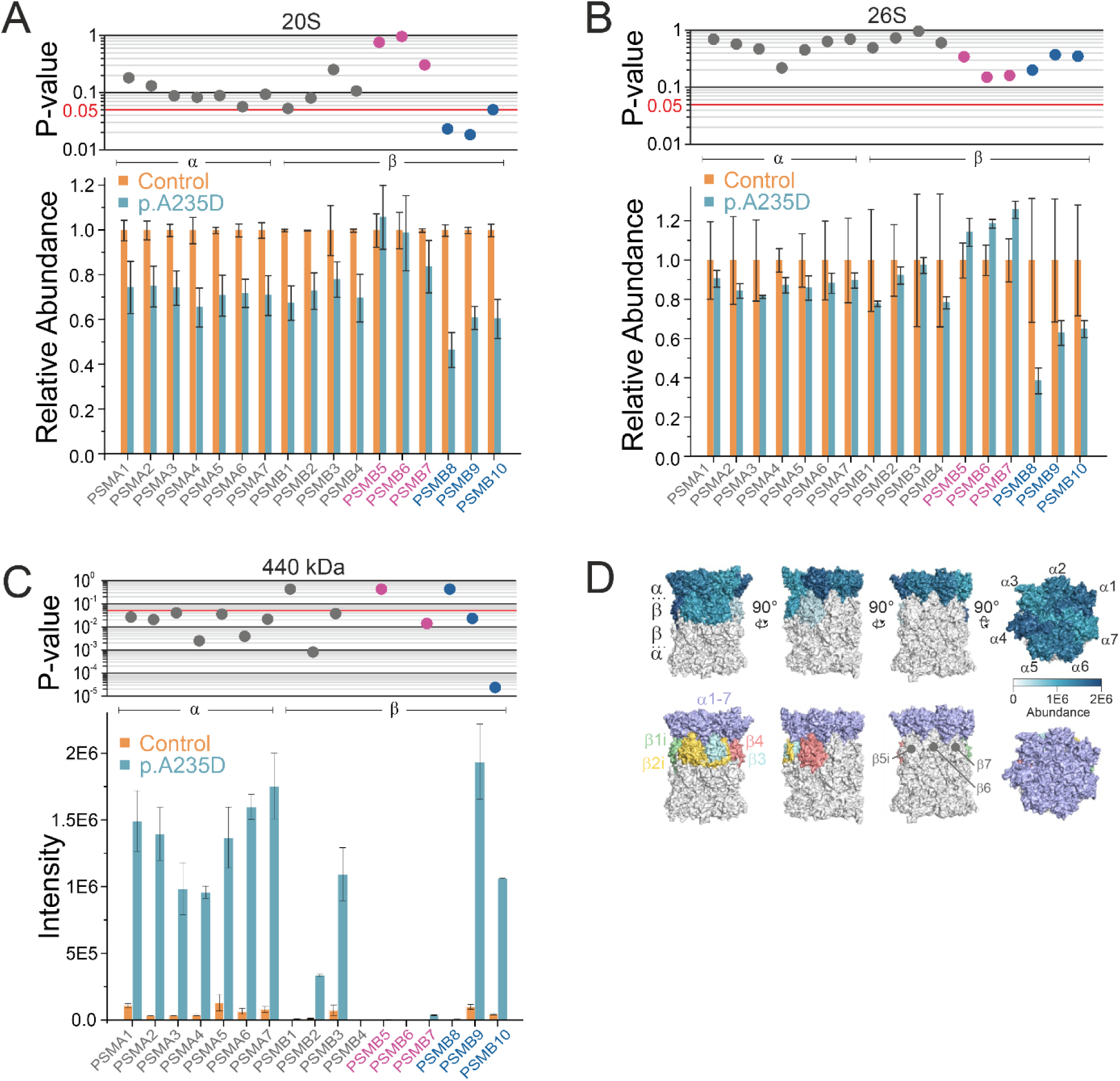
Composition of α- and β-subunits at the 20S, 26S complexes and the ∼440 kDa intermediate. **A**: Quantification of α- and β-subunits at the 20S. **B:** Quantification of α and β subunits at the 26S. **C**: Quantification of the abundance of α and β subunits at the ∼440 kDa intermediate. (A-C) Mean ± standard deviation (n=2). **D**: Upper panel: cartoon representation of the 20S IP where the upper half of the structure, comprising one α and one β ring, was colored based on the average protein intensity values observed in the patient samples (n=2). Lower panel: cartoon showing the location of the identified subunits of the subcomplex. The cryo-EM structure of the human 20S IP was used for representation of the assembly intermediate (PDB: 6E5B, (46)).

### Accumulation of assembly intermediates is specific to the IP and does not include the affected PSMB8

A peak at ∼440 kDa was observed in the profiles of the affected cell line (Figure 2A/C). This subcomplex was as abundant as the 26S complex in individual 1 (Figure 2B). Upon reviewing control fibroblasts, an intermediate at the same molecular mass could also be detected, although only at very low levels. Notably, the peak contained IP-specific subunits, while SP-specific subunits were absent, indicating that this assembly intermediate belonged exclusively to the IP (Figure 2C). By analyzing the individual abundances of all α- and β-subunits, the composition of the ∼440 kDa subassembly was inferred (Figure 3C). It contained all α-subunits and four β-subunits (PSMB2, PSMB3, PSMB9, and PSMB10), but lacked others, including PSMB8 (Figure 3D). Interestingly, the abundance of PSMB2 was observed to be lower than that of the other incorporated subunits. The migration profiles of proteasome assembly chaperones PSMG1-4 and POMP revealed additional peaks around the same mass range of the ∼440 kDa intermediate, suggesting their presence within this subcomplex (Figure 4A-C). Interestingly, a smaller peak at ∼560 kDa was also detected, containing the α- and β-subunits present in the ∼440 kDa intermediate, plus POMP and 11S regulatory proteins PSME1 and PSME2. These findings suggest that, in the ∼560 kDa complex, the assembly chaperones PSMG1-4 were replaced by the 11S heptameric complex.

**Figure 4.**
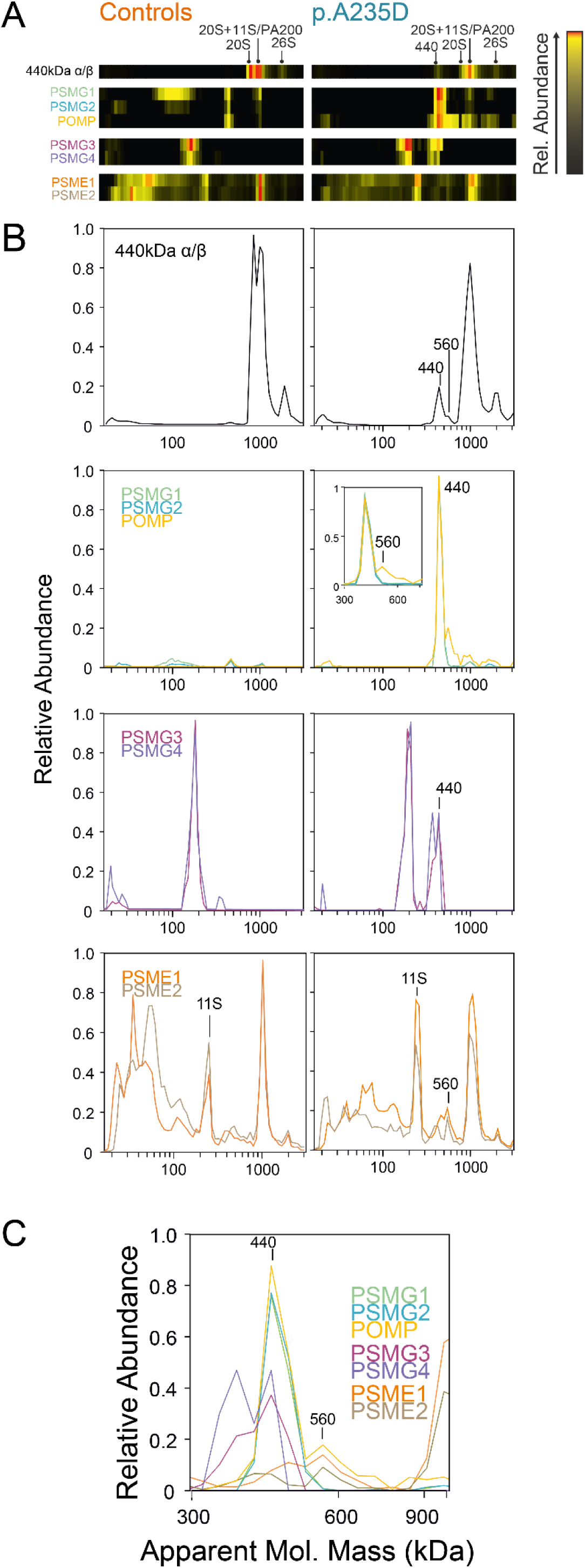
Assembly factors scaffold the accumulated IP intermediate. **A**: Heatmap representation of the average of α- and β-subunits that form the ∼440 kDa intermediate (PSMA1-7, PSMB2-3/9-10), proteasome assembly chaperones PSMG1-4 and POMP, and the components of the 11S complex, PSME1 and PSME2. Average of two independent experiments. **B**: Plots of the average migration profiles from the proteins from A. **C**: Zoom-in of the protein migration profiles around ∼440 kDa showing the stepwise incorporation and release of assembly chaperones.

To assess the maturation state of PSMB8, we analyzed the intensities of its peptides individually. Peptides from mature PSMB8 appeared majorly at the 20S and 26S peaks. However, precursor-specific N-terminal peptides were detected at low molecular mass ranges at higher levels in individual 1 compared to controls, reflecting an accumulation of free PSMB8 that remained uncleaved (Supplementary Figure 7A-C). A small peak of precursor peptides was also detected around ∼440 kDa, indicating that a very small amount of immature PSMB8 may have been transiently incorporated into the assembly intermediates. To assess the incorporation of the PSMB8 protein carrying the p.(Ala235Asp) variant into the complexes, we searched for the variant-unique peptide containing the replaced amino acid residue and analyzed its abundance distribution (Supplementary Figure 7D). The peptide carrying the variant was detected only in fibroblasts from individual 1, showing a small peak at low molecular mass ranges similar to that of the corresponding wild-type peptide, and also at the 20S mass range, but at much lower levels than the wild-type peptide. Thus, incorporation of the mutant PSMB8 protein into fully assembled proteasomes was markedly impaired, although a small fraction still integrated into the 20S complex. Furthermore, the peptide profiles of PSMB9 and PSMB10 showed accumulation at the molecular masses of the assembly intermediates, with peptides corresponding to the precursor form (Supplementary Figure 8). These findings indicate that PSMB9 and PSMB10 remained uncleaved within the ∼440 and ∼560 kDa assembly intermediates.

### Proteasome assembly factors and immune markers are dysregulated in affected fibroblasts

The differential expression of proteins in the fibroblasts was assessed, revealing a total of 113 significantly upregulated (fold-change > 3) and 99 significantly downregulated proteins (fold change < 0.33). Notably, proteasome subunits POMP and PSMG2 were upregulated in fibroblasts with the p.(Ala235Asp) variant, while other subunits remained at normal levels (Figure 5A). Furthermore, an increase in some interferon-stimulated genes and inflammatory markers was detected, including ERAP2, IFI44L, IFI44, RSAD2, and STEAP4 (Supplementary Data 2). In contrast, markers of endoplasmic reticulum stress associated with the unfolded protein response (UPR) showed comparable expression levels between cell lines. Gene Ontology enrichment analyses revealed enrichment for four terms: immune response (biological process), cell adhesion (biological process), apical plasma membrane (cellular component) and G protein-coupled receptor activity (molecular function) (Figure 5B, Supplementary Table 6).

**Figure 5.**
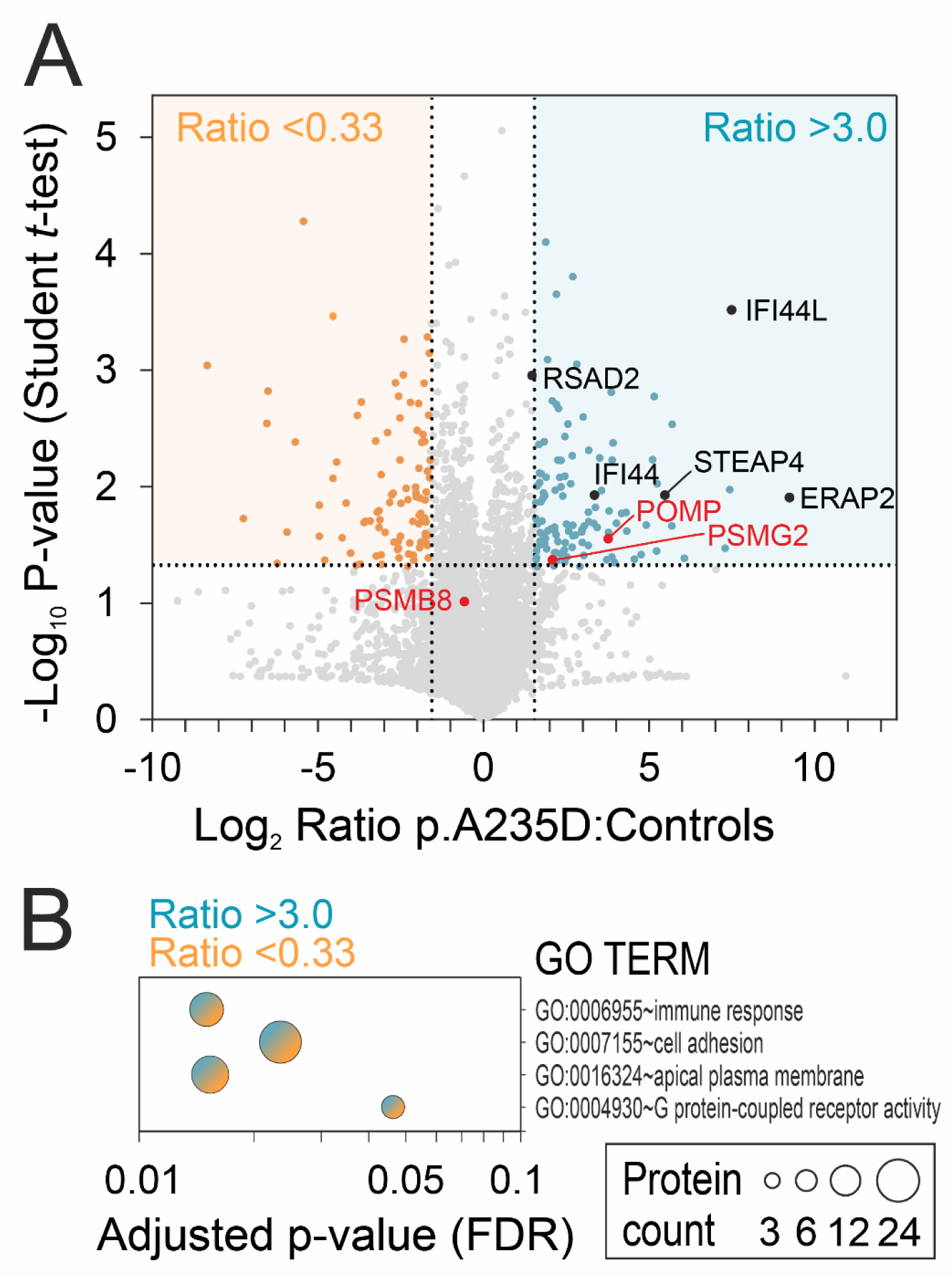
Differential expression analysis of proteins and pathway analysis. **A**: Volcano plot showing differentially expressed proteins in the p.(Ala235Asp) cell line. The orange region represents significantly downregulated proteins (fold change < 0.33), while the blue region indicates significantly upregulated proteins (fold change > 3). Differentially expressed proteins involved in proteasome assembly and PSMB8 are highlighted in red. Other highlighted inflammatory proteins are marked in black. **B**: GO-term enrichment analysis of dysregulated proteins revealed significance for four terms. Blue indicates the presence of upregulated proteins, and orange represents downregulated proteins.

## Discussion

PRAAS-related diseases were first described in association with biallelic variants in *PSMB8* and were later expanded to include pathogenic variants affecting other subunits (17–19). Although initial reports characterized recessive forms of the disease, in recent years causative monoallelic variants have been identified in two out of three IP catalytic subunits: *PSMB9* and *PSMB10* (23, 24). In this study, we describe the first individuals harboring *de novo* monoallelic variants in *PSMB8*, the remaining catalytic subunit, presenting with a PRAAS-like syndrome with immunodeficiency.

Proteasome dysfunction can result from various mechanisms, including disturbances in assembly, altered RNA or protein expression, impaired propeptide cleavage, or changes in the catalytic center. Using complexome profiling, we investigated the underlying mechanism of the p.(Ala235Asp) variant, which is located near the interface between the PSMB8 and PSMB2 subunits of opposing β-rings. This region comprises a hydrophobic patch critical for β-ring/β-ring interaction (Figure 1B). Variant p.(Ala235Asp) severely impaired the assembly of the IP but without affecting the abundance of PSMB8 protein expression, consistent with a dominant-negative mechanism. Frustration index analysis further supported this, showing an increase in highly frustrated contact pairs in the surrounding environment, suggesting a locally destabilized region that could interfere with subunit folding or interface formation.

In contrast, the p.(Ser90Phe) substitution is not located at the interface between proteasome subunits but instead affects a key loop involved in the functional dynamics of the active site (S1 pocket, Figure 1B). Frustration index analysis showed a marked increase in minimally frustrated residues around p.(Ser90Phe), suggesting excessive stabilization and rigidity in a region that normally requires conformational flexibility. This rigidity may impair proteasome function by disrupting the active site, and perturbing subunit interactions through neighboring residues potentially compromising complex assembly and stability. Interestingly, this residue lies in close proximity to previous reported *PSMB8* variants in autosomal-recessive PRAAS: p.(Ala92Thr), p.(Ala92Val), p.(Ala94Pro), p.(Met117Val) and p.(Ser118Pro) (18, 20, 21, 26–29). Notably, p.(Ser90Phe) was predicted to exert a more pronounced destabilizing effect than these recessive variants, both in terms of overall energy change and local frustration patterns. In overall, our findings suggest that p.(Ser90Phe) and p.(Ala235Asp) affect local protein stability and the surrounding interaction network more profoundly than recessive variants, which may explain their dominant pathogenic behavior.

The observed disruption in proteasome assembly in fibroblasts with the p.(Ala235Asp) variant affected the 20S and 26S complexes of the IP, but not the SP, as observed by the abundances of the catalytic subunits within these complexes. Not only was the abundance of PSMB8 reduced, but all three IP-specific subunits were present in substoichiometric amounts within the 20S and 26S assemblies. Previous studies have shown that the incorporation of IP-specific catalytic subunits is cooperative, favoring the formation of homogeneous IP assemblies over mixed proteasome forms (13, 30–32). Specifically, the incorporation of PSMB8 is strongly favored over its homolog PSMB5 in subassemblies that already contain PSMB9 and PSMB10, which are incorporated earlier in the assembly process (31, 32). In line with this, we observed an accumulation of assembly intermediates generated prior to the incorporation step of PSMB8 containing the two IP-specific catalytic subunits PSMB9 and PSMB10, along with assembly factors (6, 13, 14). A schematic representation of the assembly defect is represented in Figure 6. In this structure, PSMB2 was markedly less abundant than other subunits, and we hypothesize that the accumulated intermediate represents a mixture of subassemblies in which PSMB2 is incorporated only in a subset. The accumulation of this intermediate, together with the presence of free PSMB8, may reflect impaired incorporation of the variant protein. A weak signal corresponding to the same intermediate was also detected in control cell lines nonetheless, suggesting that this intermediate may be formed normally and is inherently more stable than others, making it more prone to accumulation in mutant cells. This suggests that incorporation of the variant PSMB8 may still occur, but the resulting structure may be unable to progress further in the assembly pathway and is subsequently targeted for degradation. The data also suggested that this incomplete assembly intermediate progressed and connected with the 11S regulatory particle. However, the complex remained catalytically inactive as propeptide-specific sequences were detected for both PSMB9 and PSMB10 at the molecular masses corresponding to the assembly intermediate (Supplementary Figure 8). This may be related to the requirement of PSMB8 for the proteolytic cleavage of their propeptides (11, 31). SP-specific subunits were expressed at similar levels across cell lines, with no evidence of compensatory upregulation. Such upregulation has been reported in PSMB8 knock-out mouse models, where, in contrast to the present case, PSMB8 protein expression is heavily downregulated or absent (30, 33, 34). Notably, among all proteasome subunits, only the expression of the assembly factors PSMG2 and POMP was significantly increased. This increase may represent a compensatory response to stalled intermediates or an effect due to their retention within the aberrant assembly intermediate, which could prevent normal turnover and degradation.

**Figure 6.**
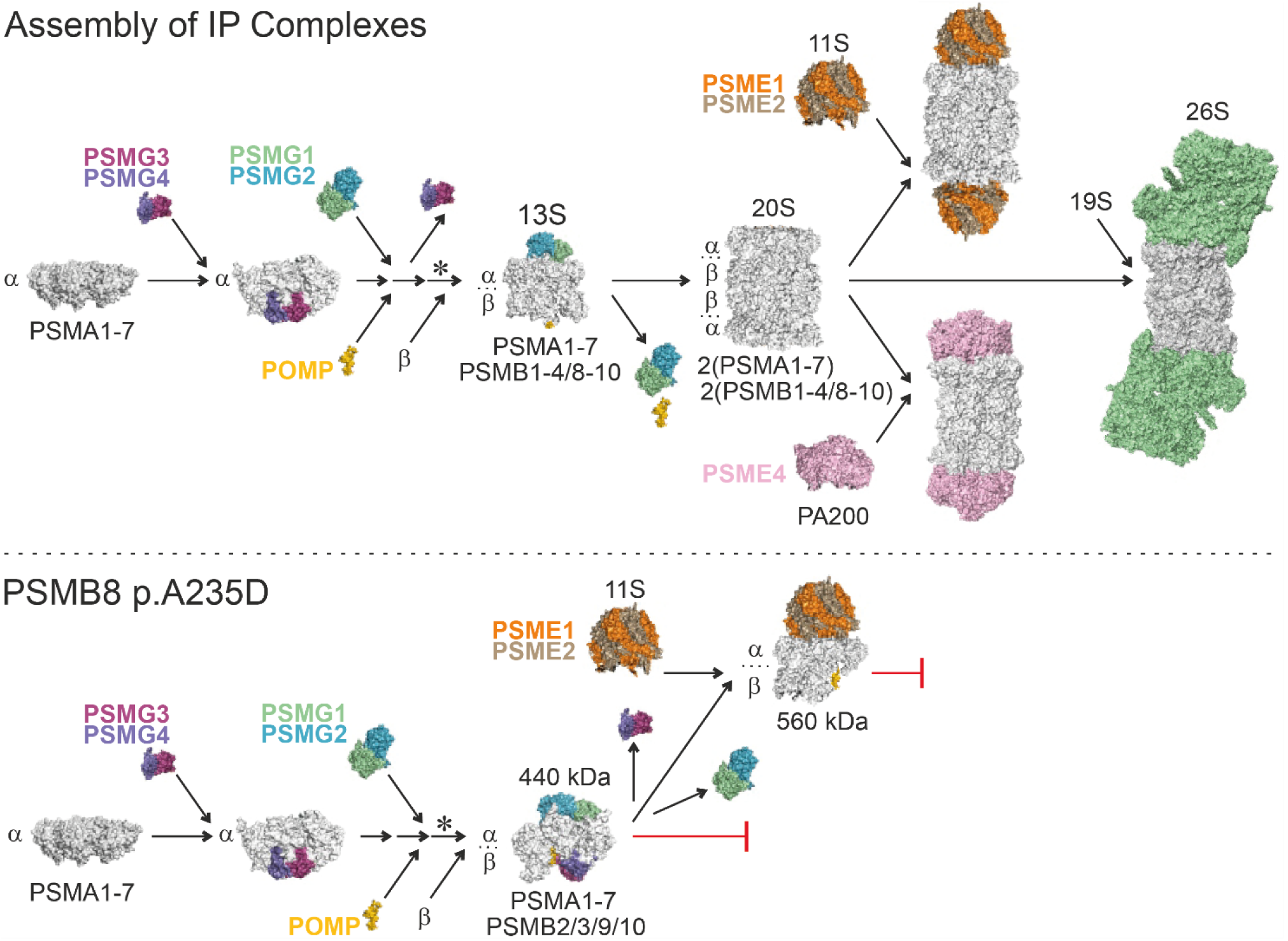
Proposed model illustrating differences in IP assembly between the control and p.(Ala235Asp) cell lines. Schematic representation of the IP assembly pathway comparing the control and p.(Ala235Asp) mutant cell lines. In the control cell line, all β subunits are efficiently incorporated into assembly intermediates, leading to the formation of the 20S core particle and, subsequently, fully assembled proteasomes containing regulatory particles*. In contrast, the p.(Ala235Asp) variant results in defective incorporation of PSMB8 and of β-subunits that are incorporated later in the assembly process. This leads to the accumulation of an aberrant ∼440 kDa assembly intermediate that still contains all assembly factors. A portion of this intermediate progresses to form a ∼560 kDa complex, in which the assembly factors, except for POMP, have been replaced by the 11S regulatory particle. This model highlights impaired proteasome biogenesis as a potential pathogenic mechanism. * β subunits are incorporated in a sequential order during proteasome assembly (according to (13)).

Monoallelic pathogenic variants in *PSMB9*, *PSMB10* and the assembly chaperone *POMP* are described to cause proteasome dysfunction by disrupting normal proteasome assembly (22–24, 35). Interestingly, all reported variants in the three catalytic subunits are missense changes, and almost all are located at the interface between the two β-rings, a region likely critical for half-mer coupling during proteasome assembly. In line with our results, Kanazawa et al. demonstrated that the p.(Gly156Asp) variant in *PSMB9* impaired the incorporation of all three catalytic subunits within the 20S and 26S IP complex, resulting in the accumulation of abnormal assembly intermediates containing immature PSMB9 and the assembly factor POMP (23). In comparison to recessive PRAAS cases, the 26S activity remained sufficient, as demonstrated by the absence of ubiquitin accumulation in skin biopsy and fibroblast cells from the mouse model (23). Variants affecting *POMP* are frameshifts that escape nonsense-mediated decay, resulting in the production of a truncated protein (22). Since POMP is a key assembly factor for all proteasome types, these variants led to deficient incorporation of both SP-specific and IP-specific subunits, as well as some α-subunits. Moreover, the 26S activity was compromised, and ubiquitin-conjugates accumulated within fibroblasts obtained from affected individuals (22).

The clinical features of the individuals described in this study show significant overlap with “PRAAS with immunodeficiency” (PRAAS-ID) caused by monoallelic variants in *PSMB9* (23, 35). These individuals exhibited systemic hyperinflammation accompanied by immunodeficiency and developed chronic lung disease with pulmonary hypertension similar to the cases described here (Table 2). Neither the cases described with variants in *PSMB9* nor those reported here presented cases showed signs of lipodystrophy or joint abnormalities, which have been reported in recessive PRAAS disorders. Immunodeficiency has also been reported in individuals harboring monoallelic variants in *PSMB10* (24), in the *PSMB10* mouse model (23, 36) and in *POMP* (22), but is rarely observed in recessive PRAAS cases. B-cell immunodeficiency with pan-hypogammaglobulinemia is a shared feature among individuals carrying monoallelic variants in any of the IP catalytic subunit. However, individuals described in this study did not also exhibit alterations in T-cell populations or function.

PSMB9 and PSMB10 are components of both the IP and the thymoproteasome, whereas PSMB8 is replaced by PSMB11 (β5t) in the thymoproteasome (6). While the IP contributes to antigen processing in T-cells, the thymoproteasome is involved in T-cell development by generating peptides for the positive selection of CD8+ T-cells during thymic maturation (37, 38). The fact that only the IP function was disturbed may partially explain why T-cell populations and functions are not affected in individuals with monoallelic *PSMB8* variants.

Individuals carrying variants in *POMP* exhibit B- and T-cell dysregulation, and additionally, they also develop autoantibodies and increased gammaglobulins, which differs from the phenotype observed in individuals with defects in the three IP-specific subunits. These differences may be related to the broader role of POMP, affecting all proteasome types.

The investigation of proteasome assembly is challenging due to the large size and complexity of its structure, involving many different proteins. In this context, we demonstrated that complexome profiling is a valuable technique to dissect the assembly process and identify variant-specific disruptions of macromolecular complexes. Such a technique is especially relevant to uncover and assess the clinical significance of missense variants in proteasome subunits, which remains particularly challenging to predict, despite advances in novel computational tools (39). Moreover, our findings support the concept that alternative inheritance patterns should be considered when missense variants occur in components of multiprotein complexes as dominant-negative effects may be involved.

To conclude, we describe two individuals with early-onset systemic autoinflammation and immunodeficiency caused by monoallelic *de novo* variants in PSMB8. Their clinical features overlap with other monoallelic PRAAS disorders and fit within the emerging description of PRAAS-ID. This report expands the spectrum of PSMB8-associated disease and broadens the clinical and molecular landscape of PRAAS.

## Materials and Methods

### Inclusion and ethical considerations

In this study, we enrolled two individuals who had previously been identified through sequencing conducted in either a diagnostic or research setting at their respective centers. Individual 1 was part of the biobank for rare diseases and was enrolled in an international research study to diagnose the undiagnosed at the Radboud University Medical Center (Radboudumc) under a protocol approved by the Institutional Review Board of CMO Radboudumc and METC East Nijmegen, the Netherlands (2018-4985 and 2019-5554).

Individual 2 was found via gene matching through the collaborative platform GeneMatcher (40). This individual was enrolled in the Undiagnosed Diseases Network (UDN) study, a multicenter study approved by the National Institutes of Health IRB (15HG0130). Both participants and/or their caregivers gave written informed consent.

### Genome sequencing and analysis

Sequencing was performed by each institution through routine exome or genome sequencing. For Individual 1, genomic DNA isolated from whole blood was collected for singleton short-read genome sequencing. Sequencing reads were mapped to the hg38 reference genome. The genome analysis included single nucleotide variants, copy number variants, structural variants, and short tandem repeats, both in the coding and noncoding regions. *In silico* predictors, along with allele frequency data from gnomAD v4.0.0, were used for a comprehensive assessment of the variants (41). Candidate variants were segregated in parental DNA with Sanger sequencing, allowing *de novo* assessment. More details are provided in the Supplementary Methods. Individual 2 underwent trio genome sequencing through the UDN Sequencing Core at Baylor Genetics as previously described (42), and research reanalysis was performed at the BCM UDN site. The research genome reanalyses prioritized rare, *de novo* and biallelic variants that had an allele frequency of < 1% in gnomAD v4.0.0 and in the BCM UDN internal sequencing database (> 1200 samples). Codified Genomics was used for variant filtering and prioritization.

The NM_148919.4 transcript of *PSMB8* is used for variant annotation and protein change reporting throughout this manuscript.

### Interferon (IFN) type I signature

For individual 1, based on the expression levels quantified from PAXgene Blood RNA tubes by RT-PCR of five IFN-related genes (IFI44, IFI44L, IFIT1, IFIT3, and MX1) and a reference gene (ABL) an interferon-type 1 (IFN-1) gene signature score was calculated for each sample, as previously described (43, 44). A value ≥ 9.4 was considered positive.

For individual 2, the IFN score calculation was adapted from (45) with the following modifications. RNA was extracted from cryopreserved peripheral blood mononuclear cells (PBMCs) using the Quick RNA Miniprep Kit (Zymo Research) per manufacturer’s instructions, and 33 ng converted to cDNA using the GoScript Reverse Transcription System (Promega). Real time PCR was performed using TaqMan Fast Advanced Master Mix and commercially available primer/probe sets (both Applied Biosystems) on a Roche LightCycler 96. The relative abundance of each target transcript was normalized to GAPDH, and data are expressed relative to a single control patient.

### Cytokine, chemokine and soluble receptor production measurements

For cytokine, chemokine and soluble receptor production measurements, serum was collected from individual 1. Concentrations (pg/mL) of human BCA-1, MIP-3, MIP-4, GM-CSF, IFNγ, IL-1β, IL-2, sIL-2Rα, IL-4, IL-5, IL-6, IL-7, IL-8, sVEGFR-2, sVCAM-1, IL-17A, IL-21, sTNFR2, IL-18, VEGF-D, IP-10, MIG, RANTES, TNFα, PF4, Endostatin, and TIMP-1 were measured according to the manufacturer’s instructions. For all factors, measured values below the lower limit of detection are represented by this lowest detection value. Serum analysis was performed using MILLIPLEX® Multiplex Assays (Merck Millipore) using a Flexmap 3D system.

### Structural biology analysis

The crystal structure of the human 20S immunoproteasome (PDB: 6E5B (46)) was used to model the variants p.(Ser90Phe) and p.(Ala235Asp) within the PSMB8 subunit. These variants were also modeled in the experimentally resolved 3D structures of the 26S proteasome homologue, using the PSMB5 subunit (PDB: 6MSB (47), 60.3% sequence identity). We estimated the changes in protein stability induced by the variants using the FoldX energy function (v.5.0) (48). Structural models for each variant were generated using RepairPDB and BuildModel functions, with five iterations of sidechain rotamer adjustments. The average differences in free energies between wildtype and the variant structures were then calculated in kcal/mol.

We performed additional analysis to account for shifts in energetic distributions in the variant environment using the 20S immunoproteasome structure. A measure known as frustration index (FI) was computed for the wildtype structure and for each of the variant models of interest using Frustratometer2 (49). The metric measures how favorable a native contact is by comparing its contribution to the overall protein energy with the energies of alternative contacts at the same site. This involves evaluating the energy of the native amino acid pair within the protein structure against the distribution of energies that would result from substituting different residues at that position (mutational FI) or altering the local environment of the interacting pair (configurational FI). We used mutational FI and configurational FI to assess variant effects, calculated for all contacting residues within 5Å of the variant site. A contact is defined as minimally frustrated if the score is >0.78, meaning the site is stable and most other amino acids at that position are unfavorable. A contact is highly frustrated if the score is <-1.00, meaning most other amino acids would be more favorable for folding at that position. Scores in-between the limits −1.00 and 0.78 are termed as neutral. Highly frustrated regions often correspond to physiologically relevant sites and typically cluster at the protein surface, while minimally frustrated regions tend to be buried and often correspond to the folded hydrophobic core of the protein (50, 51). We analyzed energetic shifts in the complex brought about by the variants p.(Ser90Phe) and p.(Ala235Asp) by comparing FI values for the wildtype and the mutants; autosomal recessive PRAAS described pathogenic variants p.(Ala92Thr), p.(Ala92Val), p.(Ala94Pro), p.(Met117Val), and p.(Ser118Pro) were also analyzed for comparison. Both polypeptide chains of PSMB8 were used for the calculations, thus a median was calculated for each variant from their single-residue frustration index values.

### Cell lines and IFNγ-stimulation

Human skin fibroblasts from individual 1 harboring the heterozygous missense mutation p.(Ala235Asp) and from two healthy controls were cultured in DMEM (Gibco) supplemented with 10% FCS (Capricorn scientific) and antibiotic/antimycotic solution in a humidified, 5% CO2 atmosphere at 37 °C. To enhance the expression of immunoproteaseome-specific subunits, cells were stimulated for 48 h with 250 U/ml human recombinant IFNγ (ImmunoTools). Cells from four 175-cm2 flasks were harvested by trypsinization, washed with PBS and centrifuged at 1000 *g* for 5 min at 4°C.

Human osteosarcoma 143B cells (cell line GM05887, Coriell Institute for Medical Research) were cultured in Dulbecco’s Modified Eagle’s Medium (DMEM, Gibco 41-965-039) supplemented with 10% fetal bovine serum (FBS-11A, Capricorn scientific) and 1% antibiotic/antimycotic solution (30-004-CL, Corning) in a humidified, 5% CO2 atmosphere at 37°C.

Human monocytic cell line THP-1 (ATCC® TIB202™) was cultured in RPMI-1640 supplemented with 10% FBS-11A, 100 U/ml penicillin, 100 µg/ml streptomycin, 250 n/ml amphotericin B and 0.05 mM β-mercaptoethanol in a humidified, 5% CO2 atmosphere at 37°C. THP-1 monocytes were differentiated with 10 ng/ml phorbol 12-myristate 13-acetate for 48 h and macrophages were harvested by trypsinization.

### Cell fractionation

Fibroblast, 143B and THP-1 cell pellets were resuspended in 4 ml ice-cold homogenization buffer (250 mM sucrose, 1 mM EDTA, 20 mM Tris/HCl, pH 7.4) and disrupted mechanically by 15 strokes, passing the cell suspension through a 20 G needle fixed to a 5-ml syringe on ice.

Fibroblasts cell homogenates were centrifuged at 1000 *g* for 10 min at 4°C and the supernatants were centrifuged at 21000 *g* for 10 min at 4°C. Pellets were resuspended in homogenization buffer and the protein concentration was determined by Lowry. Suspension aliquots containing 200 µg protein were centrifuged at 21000 *g* for 20 min at 4°C. Supernatants were discarded and the pellets were shock frozen in liquid nitrogen and stored at −80°C.

143B cell homogenates were centrifuged at 1000 *g* for 10 min at 4°C and the supernatant was centrifuged at 6000 *g* for 10 min at 4°C. Pellets were resuspended in homogenization buffer and the protein concentration was determined by Lowry. THP-1 cell homogenates were centrifuged at 1000 *g* for 10 min at 4°C and the protein concentration was determined from the supernatants.

### Blue-Native PAGE and cutting

Samples containing 200 µg of protein were thawed and resuspended at 10 mg protein/ml in solubilization buffer (0.5 M 6-aminohexanoic acid, 1 mM EDTA, 50 mM imidazole/HCl, pH 7.0) and solubilized with 6 g digitonin/g protein. Protein solutions were centrifuged at 22000 *g* for 20 min at 4°C and protein concentration of the supernatant was determined by Lowry. An aliquot containing 0.1 mg protein was mixed with 5% Coomassie blue (Serva Blue G) in 0.5 M aminohexanoic acid and loaded on a 4-16% polyacrylamide gradient gel. Proteins were separated by gel electrophoresis at 4°C as described previously (52). After electrophoresis, gels were fixed in 50% methanol, 10% acetic acid, 10 mM ammonium acetate and stained with Coomassie blue (Supplementary Figure 9A). Gels were destained in 10% acetic acid, washed with water, documented and each lane was cut into 60 even pieces and transferred to a 96-well MultiScreen-BV. 1.2 µm filter plate (Millipore).

### In-gel digestion

Gel pieces were destained for 30 min in 50% methanol, 50 mM ammonium bicarbonate (ABC) at room temperature and centrifuged at 600 *g* for 2 min. This step was repeated 3-4 times until the Coomassie blue dye was removed entirely. Then, gel pieces were incubated for 1 h in 5 mM dithiothreitol, 50 mM ABC followed by 45 min incubation with 15 mM 2-chloroacetamide, 50 mM ABC. After washing once with 50% methanol, 50 mM ABC, gel pieces were let dry at room temperature for 30 min and rehydrated with 20 µl of 5 µg trypsin/ml in 50 mM ABC for 30 min at 4°C. After adding 50 µl 50 mM ABC, plates were sealed and incubated overnight at 37°C. Peptide solutions were transferred to a PCR plate by centrifugation at 600 *g* for 2 min. The gel pieces were washed once with 50% acetonitrile, 5% formic acid and the filtrates were collected into the same PCR plate. Peptide solutions were dried for 2.5 h at 45°C in a centrifuge concentrator (Concentrator plus, Eppendorf). Peptides were dissolved in 20 µl 0.1% formic acid.

### LC-MS/MS

From fibroblasts samples, 2 µl were injected and subjected to liquid chromatography tandem mass spectrometry (LC-MS/MS) on a quadrupole-orbitrap hybrid orbitrap mass spectrometer (Exploris 480, Thermo Fisher Scientific) coupled at the front end to an ultra-high pressure liquid chromatography system (Vanquish neo UHPLC System, Thermo Fisher). Attached to the UHPLC was a peptide trap (100 µm x 20 mm, 100 Å pore size, 5 µm particle size, C18, Nano Viper, Thermo Fisher) for online desalting and purification, followed by a 25 cm C18 reversed-phase column (75 µm x 250 mm, 130 Å pore size, 1.7 µm particle size, peptide BEH C18, nanoEase, Waters). Peptides were separated using a 35 min method with linearly increasing ACN concentration from 2% to 30% ACN over 25 minutes.

MS/MS measurements from fibroblasts samples were performed on a quadrupole-orbitrap hybrid mass spectrometer (Exploris 480, Thermo Fisher Scientific). Eluting peptides were ionized using a nano-electrospray ionization source (nano-ESI) with a spray voltage of 1,800 V and analyzed in data-independent acquisition (DIA) mode. For each MS1 scan, ions were accumulated for a maximum of 240 ms or until a charge density of 3 x 10^6^ ions (AGC Target) was reached. Fourier-transformation based mass analysis of the data from the orbitrap mass analyzer was performed covering a mass range of m/z 400 – 1,400 with a resolution of 120,000 at m/z 200. Within a precursor mass range of m/z 380-980 fragmentation in DIA-mode with m/z 12 isolation windows and m/z 1 window overlaps was performed.

Fragmentation was performed at normalized collision energy of 28% using higher energy collisional dissociation (HCD). An AGC target of 2 x 10^6^ ions or a maximum of 54 ms was set. Orbitrap resolution was set to 30,000 with a scan range from m/z 350-2000.

From 143B and THP-1 samples, 5 µl of the peptide solutions were analyzed by liquid chromatography electrospray ionization tandem mass spectrometry (LC-ESI-MS/MS) in a Q-Exactive mass spectrometer (Thermo Fisher Scientific) equipped with an Easy nLC1000 nano-flow high-performance liquid chromatography system at the front end. Peptide separation was performed with an emitter column (15cm L x 100 µm ID x 360 µm OD x 15 µm orifice; MSWil, CoAnn Technologies, LLC) filled with ReproSil-Pur C18-AQ reverse phase beads (3 µm particle size, 120 Å pore size; Dr. Maisch GmbH) using a 30 min linear gradient of 5 to 35% acetonitrile with 0.1% formic acid. The mass spectrometer operated in positive ion switching automatically between MS and data dependent MS/MS, fragmenting the twenty most intense ions per precursor scan. Full scan MS mode (400 to 1400 m/z) was operated with automatic gain control target of 1 × 10^6^ ions, 70000 resolution and a maximum ion transfer of 20 ms. Selected ions for MS/MS were analyzed using the following parameters: resolution 17,500; AGC target of 1 × 10^5^; maximum ion transfer of 50 ms; 4.0 m/z isolation window and dynamic exclusion of 30.0 s was used.

### Proteomics data analysis

LC-MS/MS data from fibroblasts were searched with the CHIMERYS DIA algorithm integrated into the Proteome Discoverer software (v3.1.0.638, Thermo Fisher Scientific) against a reviewed human Swissprot database (obtained November 2023) using Inferys 3.0 fragmentation as prediction model. Carbamidomethylation was set as a fixed modification for cysteine residues. The oxidation of methionine was allowed as a variable modification. A maximum number of one missing tryptic cleavage was set. Peptides between 7 and 30 amino acids were considered. A strict cutoff (FDR < 0.01) was set for peptide identification. Quantification was performed by CHIMERYS based on fragment ions. The mass spectrometry proteomics data have been deposited to the ProteomeXchange Consortium via the PRIDE (53) partner repository with the dataset identifier PXD064505.

A new database search was performed to identify the PSMB8 variant A235D. To this end, the raw data were searched with the DIA-NN algorithm (Version 1.9.1 (54)) against a human database (obtained November 2023) which included the PSMB8 variant A235D. The oxidation of methionine, the N-terminal methionine excision, and the acetylation of the protein N-terminus were allowed as variable modifications. A maximum number of two missing tryptic cleavages was set. Peptides between 7 and 30 amino acids were considered. A strict cutoff (FDR < 0.01) was set for peptide identification in double pass mode. Match between runs was activated.

LC-MS/MS raw files from 143B and THP-1 samples were analyzed using MaxQuant 1.5.0.25, and 1.6.17.0, respectively. 143B spectra were matched against the human NCBI Reference Sequence Database release 55 with reverse decoy and a false discovery rate of 0.01. THP-1 spectra were marched against the Uniprot database of canonical isoforms downloaded on May 2021.

Protein groups text files containing the abundance values of each protein across all gel fractions were subjected to hierarchical clustering analysis by uncentered Pearson correlation with average linkage using Cluster 3.0 (55). Protein migration profiles were visualized using NOVA v0.5.7 (56) and the differences of the protein migration profiles between controls and patient fibroblasts were scored by Hausdorff distance calculations using COPAL as described previously (57). A set of soluble and membrane protein complexes with established molecular mass and stoichiometry were used as standards to estimate the molecular masses of globular (hydrophilic) and transmembrane (hydrophobic) protein complexes in each gel fraction, respectively (Supplementary Figure 9B).

### Data availability

Mass spectrometry data from IFN**γ**-stimulated fibroblasts is available in the ProteomeXchange Consortium via the PRIDE partner repository (53) with dataset identifier PXD064505. The complexome profiling dataset is available at the Complexome Profiling Data Resource (CEDAR) (58) with identifier number CRX49.

### Statistics

Protein complex quantifications from complexome profiling data were calculated as the mean protein abundance of fibroblasts from two healthy controls and from two replicate cultures from individual 1. Error bars represent the standard deviation. The sums of all protein abundance values across the 60 fractions were used to calculate the total protein intensities. Students *t*-test, two sided, was used to analyze the differences in total protein intensities between fibroblasts from controls and from individual 1. Graph Prism 6 was used to visualize the proteomics volcano plot.

## Acknowledgements

We would like to thank Wim A. Dik and Sandra J. Posthumus-van Sluijs for performing the type I interferon signature analysis, and Esther van Rijssen and Evelien Sprenkeler for performing the Luminex analyses. We also thank Laura Lubbers for drawing patient samples. This work was supported by Metakids, the United for Metabolic Diseases consortium (UMD) under grant number UMD-ZOE-2022-012, and the National Institute of Neurological Disorders and Stroke of the National Institutes of Health under Award Numbers U01HG007709 and U01HG007942. The content is solely the responsibility of the authors and does not necessarily represent the official views of the National Institutes of Health. This study was also supported by grants from the Deutsche Forschungsgemeinschaft (DFG; INST 337/15-1, INST 337/16-1, INST 152/837-1 and INST 152/947-1). We thank the Core Facility Mass Spectrometric Proteomics as part of the Technology Platform Mass Spectrometry (TPMS) at University of Hamburg (UHH) and University Medical Center Hamburg-Eppendorf (UKE) for support with mass spectrometric measurements and analysis funded by the Deutsche Forschungsgemeinschaft (DFG, German Research Foundation) 518551069.

## Author contributions

R.W., C.I.v.d.M., S.B.W., R.A.W., M.M.O., and S.G.-C. contributed to the conception and design of the study. R.W., M.W.-H and M.M.O. performed the genetic data analysis. R.W., C.I.v.d.M., S.K.U., J.A.R., T.P.V. and S.K.N. collected the clinical data. R.W., T.P.V., A.C.O., B.S., T.M., H.S., R.v.B. and S.G.-C. performed the experimental work. G.R. performed the structural biology analysis. R.L.S. performed the cytokine analysis. S.B.W., R.A.W., M.M.O., C.v.K., L.E.L.M.V. and A.H. supervised the project. R.W. and S.G.-C. wrote the initial draft of the manuscript. S.B.W., R.A.W. and M.M.O. provided critical feedback and assisted with manuscript revisions. All authors read and approved the final version of the manuscript.

## Consortia

### Undiagnosed Diseases Network

Jill A. Rosenfeld^5^, Tiphanie P. Vogel^6^, Monika Weisz-Hubshman^5,8^A full list of members appears in the Supplementary Information.

## References

1. Rousseau A, Bertolotti A. Regulation of proteasome assembly and activity in health and disease. Nat Rev Mol Cell Biol. 2018;19(11):697–712.

2. Murata S, Yashiroda H, Tanaka K. Molecular mechanisms of proteasome assembly. Nat Rev Mol Cell Biol. 2009;10(2):104–15.

3. Kaneko T, Hamazaki J, Iemura S, Sasaki K, Furuyama K, Natsume T, et al. Assembly pathway of the Mammalian proteasome base subcomplex is mediated by multiple specific chaperones. Cell. 2009;137(5):914–25.

4. Tanaka K. The proteasome: overview of structure and functions. Proc Jpn Acad Ser B Phys Biol Sci. 2009;85(1):12–36.

5. Arendt CS, Hochstrasser M. Identification of the yeast 20S proteasome catalytic centers and subunit interactions required for active-site formation. Proc Natl Acad Sci U S A. 1997;94(14):7156–61.

6. Murata S, Takahama Y, Kasahara M, Tanaka K. The immunoproteasome and thymoproteasome: functions, evolution and human disease. Nat Immunol. 2018;19(9):923–31.

7. Heink S, Ludwig D, Kloetzel PM, Kruger E. IFN-gamma-induced immune adaptation of the proteasome system is an accelerated and transient response. Proc Natl Acad Sci U S A. 2005;102(26):9241–6.

8. Rock KL, Goldberg AL. Degradation of cell proteins and the generation of MHC class I-presented peptides. Annu Rev Immunol. 1999;17:739–79.

9. Sijts EJ, Kloetzel PM. The role of the proteasome in the generation of MHC class I ligands and immune responses. Cell Mol Life Sci. 2011;68(9):1491–502.

10. Kincaid EZ, Che JW, York I, Escobar H, Reyes-Vargas E, Delgado JC, et al. Mice completely lacking immunoproteasomes show major changes in antigen presentation. Nat Immunol. 2011;13(2):129–35.

11. Ferrington DA, Gregerson DS. Immunoproteasomes: structure, function, and antigen presentation. Prog Mol Biol Transl Sci. 2012;109:75–112.

12. Adolf F, Du J, Goodall EA, Walsh RM, Jr., Rawson S, von Gronau S, et al. Visualizing chaperone-mediated multistep assembly of the human 20S proteasome. Nat Struct Mol Biol. 2024;31(8):1176–88.

13. Watanabe A, Yashiroda H, Ishihara S, Lo M, Murata S. The Molecular Mechanisms Governing the Assembly of the Immuno- and Thymoproteasomes in the Presence of Constitutive Proteasomes. Cells. 2022;11(9).

14. Nandi D, Woodward E, Ginsburg DB, Monaco JJ. Intermediates in the formation of mouse 20S proteasomes: implications for the assembly of precursor beta subunits. EMBO J. 1997;16(17):5363–75.

15. Mendonca LO, Fremond ML. Interferonopathies: From concept to clinical practice. Best Pract Res Clin Rheumatol. 2024;38(3):101975.

16. de Jesus AA, Hou Y, Brooks S, Malle L, Biancotto A, Huang Y, et al. Distinct interferon signatures and cytokine patterns define additional systemic autoinflammatory diseases. J Clin Invest. 2020;130(4):1669–82.

17. Agarwal AK, Xing C, DeMartino GN, Mizrachi D, Hernandez MD, Sousa AB, et al. PSMB8 encoding the beta5i proteasome subunit is mutated in joint contractures, muscle atrophy, microcytic anemia, and panniculitis-induced lipodystrophy syndrome. Am J Hum Genet. 2010;87(6):866–72.

18. de Jesus AA, Brehm A, VanTries R, Pillet P, Parentelli AS, Montealegre Sanchez GA, et al. Novel proteasome assembly chaperone mutations in PSMG2/PAC2 cause the autoinflammatory interferonopathy CANDLE/PRAAS4. J Allergy Clin Immunol. 2019;143(5):1939–43 e8.

19. Sarrabay G, Mechin D, Salhi A, Boursier G, Rittore C, Crow Y, et al. PSMB10, the last immunoproteasome gene missing for PRAAS. J Allergy Clin Immunol. 2020;145(3):1015–7 e6.

20. Brehm A, Liu Y, Sheikh A, Marrero B, Omoyinmi E, Zhou Q, et al. Additive loss-of-function proteasome subunit mutations in CANDLE/PRAAS patients promote type I IFN production. J Clin Invest. 2015;125(11):4196–211.

21. Papendorf JJ, Ebstein F, Alehashemi S, Piotto DGP, Kozlova A, Terreri MT, et al. Identification of eight novel proteasome variants in five unrelated cases of proteasome-associated autoinflammatory syndromes (PRAAS). Front Immunol. 2023;14:1190104.

22. Poli MC, Ebstein F, Nicholas SK, de Guzman MM, Forbes LR, Chinn IK, et al. Heterozygous Truncating Variants in POMP Escape Nonsense-Mediated Decay and Cause a Unique Immune Dysregulatory Syndrome. Am J Hum Genet. 2018;102(6):1126–42.

23. Kanazawa N, Hemmi H, Kinjo N, Ohnishi H, Hamazaki J, Mishima H, et al. Heterozygous missense variant of the proteasome subunit beta-type 9 causes neonatal-onset autoinflammation and immunodeficiency. Nat Commun. 2021;12(1):6819.

24. van der Made CI, Kersten S, Chorin O, Engelhardt KR, Ramakrishnan G, Griffin H, et al. Expanding the PRAAS spectrum: De novo mutations of immunoproteasome subunit beta-type 10 in six infants with SCID-Omenn syndrome. Am J Hum Genet. 2024;111(4):791–804.

25. Unno M, Mizushima T, Morimoto Y, Tomisugi Y, Tanaka K, Yasuoka N, et al. The structure of the mammalian 20S proteasome at 2.75 A resolution. Structure. 2002;10(5):609–18.

26. Boyadzhiev M, Marinov L, Boyadzhiev V, Iotova V, Aksentijevich I, Hambleton S. Disease course and treatment effects of a JAK inhibitor in a patient with CANDLE syndrome. Pediatr Rheumatol Online J. 2019;17(1):19.

27. Cavalcante MP, Brunelli JB, Miranda CC, Novak GV, Malle L, Aikawa NE, et al. CANDLE syndrome: chronic atypical neutrophilic dermatosis with lipodystrophy and elevated temperature-a rare case with a novel mutation. Eur J Pediatr. 2016;175(5):735–40.

28. Patel PN, Hunt R, Pettigrew ZJ, Shirley JB, Vogel TP, de Guzman MM. Successful treatment of chronic atypical neutrophilic dermatosis with lipodystrophy and elevated temperature (CANDLE) syndrome with tofacitinib. Pediatr Dermatol. 2021;38(2):528–9.

29. Kluk J, Rustin M, Brogan PA, Omoyinmi E, Rowczenio DM, Willcocks LC, et al. Chronic atypical neutrophilic dermatosis with lipodystrophy and elevated temperature syndrome: a report of a novel mutation and review of the literature. Br J Dermatol. 2014;170(1):215–7.

30. Cetin G, Studencka-Turski M, Venz S, Schormann E, Junker H, Hammer E, et al. Immunoproteasomes control activation of innate immune signaling and microglial function. Front Immunol. 2022;13:982786.

31. Griffin TA, Nandi D, Cruz M, Fehling HJ, Kaer LV, Monaco JJ, et al. Immunoproteasome assembly: cooperative incorporation of interferon gamma (IFN-gamma)-inducible subunits. J Exp Med. 1998;187(1):97–104.

32. De M, Jayarapu K, Elenich L, Monaco JJ, Colbert RA, Griffin TA. Beta 2 subunit propeptides influence cooperative proteasome assembly. J Biol Chem. 2003;278(8):6153–9.

33. Hewing B, Ludwig A, Dan C, Potzsch M, Hannemann C, Petry A, et al. Immunoproteasome subunit ss5i/LMP7-deficiency in atherosclerosis. Sci Rep. 2017;7(1):13342.

34. de Freitas Chama LL, Ebstein F, Wiesrecker B, Wagh PR, Hammer E, Weiss FU, et al. Immunoproteasome impairment via beta5i/LMP7-deletion leads to sustained pancreatic injury from experimental pancreatitis. J Cell Mol Med. 2021;25(14):6786–99.

35. Kataoka S, Kawashima N, Okuno Y, Muramatsu H, Miwata S, Narita K, et al. Successful treatment of a novel type I interferonopathy due to a de novo PSMB9 gene mutation with a Janus kinase inhibitor. J Allergy Clin Immunol. 2021;148(2):639–44.

36. Treise I, Huber EM, Klein-Rodewald T, Heinemeyer W, Grassmann SA, Basler M, et al. Defective immuno- and thymoproteasome assembly causes severe immunodeficiency. Sci Rep. 2018;8(1):5975.

37. Nitta T, Murata S, Sasaki K, Fujii H, Ripen AM, Ishimaru N, et al. Thymoproteasome shapes immunocompetent repertoire of CD8+ T cells. Immunity. 2010;32(1):29–40.

38. Takahama Y. The thymoproteasome in shaping the CD8(+) T-cell repertoire. Curr Opin Immunol. 2023;83:102336.

39. Cheng J, Novati G, Pan J, Bycroft C, Zemgulyte A, Applebaum T, et al. Accurate proteome-wide missense variant effect prediction with AlphaMissense. Science. 2023;381(6664):eadg7492.

40. Sobreira N, Schiettecatte F, Valle D, Hamosh A. GeneMatcher: a matching tool for connecting investigators with an interest in the same gene. Hum Mutat. 2015;36(10):928–30.

41. Chen S, Francioli LC, Goodrich JK, Collins RL, Kanai M, Wang Q, et al. A genomic mutational constraint map using variation in 76,156 human genomes. Nature. 2024;625(7993):92–100.

42. Keehan L, Jiang MM, Li X, Marom R, Dai H, Murdock D, et al. A novel de novo intronic variant in ITPR1 causes Gillespie syndrome. Am J Med Genet A. 2021;185(8):2315–24.

43. Huijser E, Bodewes ILA, Lourens MS, van Helden-Meeuwsen CG, van den Bosch TPP, Grashof DGB, et al. Hyperresponsive cytosolic DNA-sensing pathway in monocytes from primary Sjogren’s syndrome. Rheumatology (Oxford). 2022;61(8):3491–6.

44. Bodewes ILA, Al-Ali S, van Helden-Meeuwsen CG, Maria NI, Tarn J, Lendrem DW, et al. Systemic interferon type I and type II signatures in primary Sjogren’s syndrome reveal differences in biological disease activity. Rheumatology (Oxford). 2018;57(5):921–30.

45. Rice GI, Forte GM, Szynkiewicz M, Chase DS, Aeby A, Abdel-Hamid MS, et al. Assessment of interferon-related biomarkers in Aicardi-Goutieres syndrome associated with mutations in TREX1, RNASEH2A, RNASEH2B, RNASEH2C, SAMHD1, and ADAR: a case-control study. Lancet Neurol. 2013;12(12):1159–69.

46. Ladi E, Everett C, Stivala CE, Daniels BE, Durk MR, Harris SF, et al. Design and Evaluation of Highly Selective Human Immunoproteasome Inhibitors Reveal a Compensatory Process That Preserves Immune Cell Viability. J Med Chem. 2019;62(15):7032–41.

47. Dong Y, Zhang S, Wu Z, Li X, Wang WL, Zhu Y, et al. Cryo-EM structures and dynamics of substrate-engaged human 26S proteasome. Nature. 2019;565(7737):49–55.

48. Schymkowitz J, Borg J, Stricher F, Nys R, Rousseau F, Serrano L. The FoldX web server: an online force field. Nucleic Acids Res. 2005;33(Web Server issue):W382-8.

49. Parra RG, Schafer NP, Radusky LG, Tsai MY, Guzovsky AB, Wolynes PG, et al. Protein Frustratometer 2: a tool to localize energetic frustration in protein molecules, now with electrostatics. Nucleic Acids Res. 2016;44(W1):W356–60.

50. Ferreiro DU, Hegler JA, Komives EA, Wolynes PG. Localizing frustration in native proteins and protein assemblies. Proc Natl Acad Sci U S A. 2007;104(50):19819–24.

51. Ferreiro DU, Hegler JA, Komives EA, Wolynes PG. On the role of frustration in the energy landscapes of allosteric proteins. Proc Natl Acad Sci U S A. 2011;108(9):3499–503.

52. Wittig I, Braun HP, Schägger H. Blue native PAGE. Nat Protoc. 2006;1(1):418–28.

53. Perez-Riverol Y, Bandla C, Kundu DJ, Kamatchinathan S, Bai JW, Hewapathirana S, et al. The PRIDE database at 20 years: 2025 update. Nucleic Acids Research. 2024;53(D1):D543–D53.

54. Demichev V, Messner CB, Vernardis SI, Lilley KS, Ralser M. DIA-NN: neural networks and interference correction enable deep proteome coverage in high throughput. Nat Methods. 2020;17(1):41–4.

55. de Hoon MJ, Imoto S, Nolan J, Miyano S. Open source clustering software. Bioinformatics. 2004;20(9):1453–4.

56. Giese KP, Aziz W, Kraev I, Stewart MG. Generation of multi-innervated dendritic spines as a novel mechanism of long-term memory formation. Neurobiol Learn Mem. 2015;124:48–51.

57. Van Strien J, Guerrero-Castillo S, Chatzispyrou IA, Houtkooper RH, Brandt U, Huynen MA. COmplexome Profiling ALignment (COPAL) reveals remodeling of mitochondrial protein complexes in Barth syndrome. Bioinformatics. 2019;35(17):3083–91.

58. van Strien J, Haupt A, Schulte U, Braun HP, Cabrera-Orefice A, Choudhary JS, et al. CEDAR, an online resource for the reporting and exploration of complexome profiling data. Biochim Biophys Acta Bioenerg. 2021;1862(7):148411.

59. Meinhardt A, Ramos PC, Dohmen RJ, Lucas N, Lee-Kirsch MA, Becker B, et al. Curative Treatment of POMP-Related Autoinflammation and Immune Dysregulation (PRAID) by Hematopoietic Stem Cell Transplantation. J Clin Immunol. 2021;41(7):1664–7.

60. Arima K, Kinoshita A, Mishima H, Kanazawa N, Kaneko T, Mizushima T, et al. Proteasome assembly defect due to a proteasome subunit beta type 8 (PSMB8) mutation causes the autoinflammatory disorder, Nakajo-Nishimura syndrome. Proc Natl Acad Sci U S A. 2011;108(36):14914–9.

61. Liu Y, Ramot Y, Torrelo A, Paller AS, Si N, Babay S, et al. Mutations in proteasome subunit beta type 8 cause chronic atypical neutrophilic dermatosis with lipodystrophy and elevated temperature with evidence of genetic and phenotypic heterogeneity. Arthritis Rheum. 2012;64(3):895–907.

62. Verhoeven D, Schonenberg-Meinema D, Ebstein F, Papendorf JJ, Baars PA, van Leeuwen EMM, et al. Hematopoietic stem cell transplantation in a patient with proteasome-associated autoinflammatory syndrome (PRAAS). J Allergy Clin Immunol. 2022;149(3):1120–7 e8.

63. Huang X, Luan B, Wu J, Shi Y. An atomic structure of the human 26S proteasome. Nat Struct Mol Biol. 2016;23(9):778–85.

