## Supplementary Information for "Complexome profiling showed impaired immunoproteasome assembly in a novel PRAAS subtype caused by monoallelic PSMB8 variants"

‡ A list of authors and their affiliations appears at the end of the paper

### Supplementary Methods

#### Genome sequencing and variant analysis

Individual 1

Genome sequencing for individual 1 was outsourced to the Beijing Genomics Institute (BGI) and performed on a BGISeq500 platform. Paired-end 100 bp reads were generated, achieving a median coverage of 30-fold. Data processing was carried out at the Radboud University Medical Center. Reads were aligned to the GRCh38 reference genome using Bwa-mem2 v2.2.1 (1), and quality control was performed with Qualimap v2.2.1 (2). Single nucleotide variants (SNVs) were called using GATK HaplotypeCaller v3.8 (3). Structural variants (SVs) were detected using Manta v1.1.0 (Illumina) (4), and copy number variants (CNVs) were identified with Canvas v1.40.0 (Illumina) (5). Short tandem repeats (STRs) were analyzed using ExpansionHunter v3.1.2 with default parameters (6). All variant types (SNVs, SVs, CNVs) were annotated using an in-house pipeline. Rare candidate SNVs were prioritized based on a gnomAD v3.1 and internal frequency of <1%. CNVs and SVs were filtered using a <1% frequency threshold in the 1000 Genomes database and an internal reference set, requiring a minimum reciprocal overlap of 90%. Additional annotations, including CADD scores, SpliceAI, phyloP, and AlphaMissense, along with the patient's phenotype, were used to prioritize potentially disease-causing variants.

#### Supplementary Results

##### Additional proteasome assemblies detected in 143B, THP-1, and control fibroblast cell lines by complexome profiling

To get insight into the whole proteasome assembly process, additional patterns observed in the migration profiles and heatmaps were assessed.

In the complexome profiling data, we observed an intermediate of the 20S proteasome assembly in THP-1 and stimulated fibroblast cell lines at very low abundances. This structure contained a complete α-ring and included subunits PSMB2, PSMB3, PSMB9 and PSMB10. The presence of immunoproteasome (IP)-specific subunits PSMB9-10, and the absence of standard proteasome (SP)-specific subunitsPSMB5–7, indicated that this intermediate is specific to the IP. The 20S assembly factors POMP and PSMG1–2 were also detected within this intermediate, consistent with their known role early in proteasome biogenesis and their dissociation later in the assembly process. The abundance of this intermediate was substantially lower than that of the mature 20S and 26S complexes, and it was not detected in the 143B cell line, which is in line with the generally low expression of the IP in this non-immunological cell type.

Regarding the 19S regulatory particles, three distinct subassemblies were detected. Two of these corresponded to base subcomplexes: one composed of PSMC3, PSMC6, and PSMD9, and the other formed by PSMC1, PSMC2, PSMD2, and PSMD5. The third intermediate was part of the 19S lid and comprised subunits PSMD3, PSMD6, PSMD7, PSMD11, PSMD12, PSMD13 and PSMD14. Migration profiles of subunits PSMC1-6 and PSMD2 also showed another peak slightly smaller than the 26S, indicating that these subunits of the base were associated first to the 20S core particle. The complete 19S structure was only observed as part of the assembled 26S proteasome complex. This complex contained the PSMC1-6 subunits forming the ATPase module, as well as non-ATPase subunits previously identified in the subassemblies, along with PSMD1, PSMD4, and PSMD8. Notably, PSMD5, PSMD9 and PSMD10 were absent from the fully assembled 26S complex, consistent with their known roles as assembly chaperones rather than structural components. The only subunit known to be part of the human 26S complex (PDB: 6MSB (17)) that was not identified by LC-MS/MS was SEM1. This protein was not efficiently detected by mass spectrometry due to its small size and the lack of tryptic peptides of a detectable length.

Finally, we also detected subassembly structures of the 11S regulatory particles. At a similar molecular mass, we observed the presence of PSME1, PSME2, and PSME3. These signals corresponded to two distinct 11S complexes: the 11S αβ heteroheptamer formed by PSME1 and PSME2, and the 11S γ homoheptamer formed by PSME3. At the same molecular mass, we also detected the PA200 regulatory particle, formed by a single molecule of PSME4. Both the 11S αβ complex and PA200 were also observed in association with the 20S core particle, alone or as part of hybrid proteasomes incorporating the 20S core particle with the 19S and either 11S αβ or PA200. In contrast, the 11S γ complex was not detected in association with the 20S core particle.

All proteasome structures identified in this study through complexome profiling are presented in Supplementary Figure 6B and summarized in Supplementary Data 1.

#### Supplementary Figures

##### Supplementary Figure 1. Interferon signature score plot for individual 2

Blue are interferon (IFN) scores from healthy controls and red is the IFN score from individual 2, from a post-lung transplant specimen (pre-transplant samples were not available). Solid line represents the mean of healthy controls, dotted lines represent 2 standard deviations from the mean.

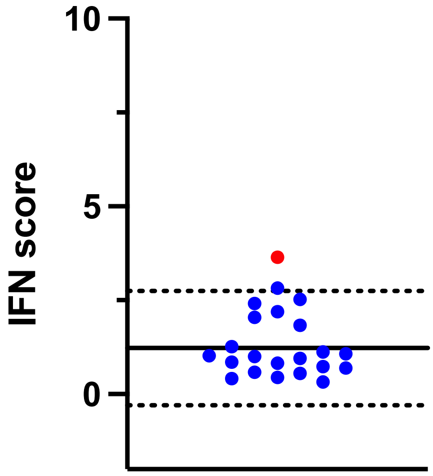

**Supplementary Figure 2. Genetic evidence of identified variants**

**A:** Visualization of the variant in genome sequencing data and Sanger sequencing traces confirming the variant in the index case and parents for family 1. **B:** Visualization of the variant in genome sequencing data for family 2, showing proband, father and mother.

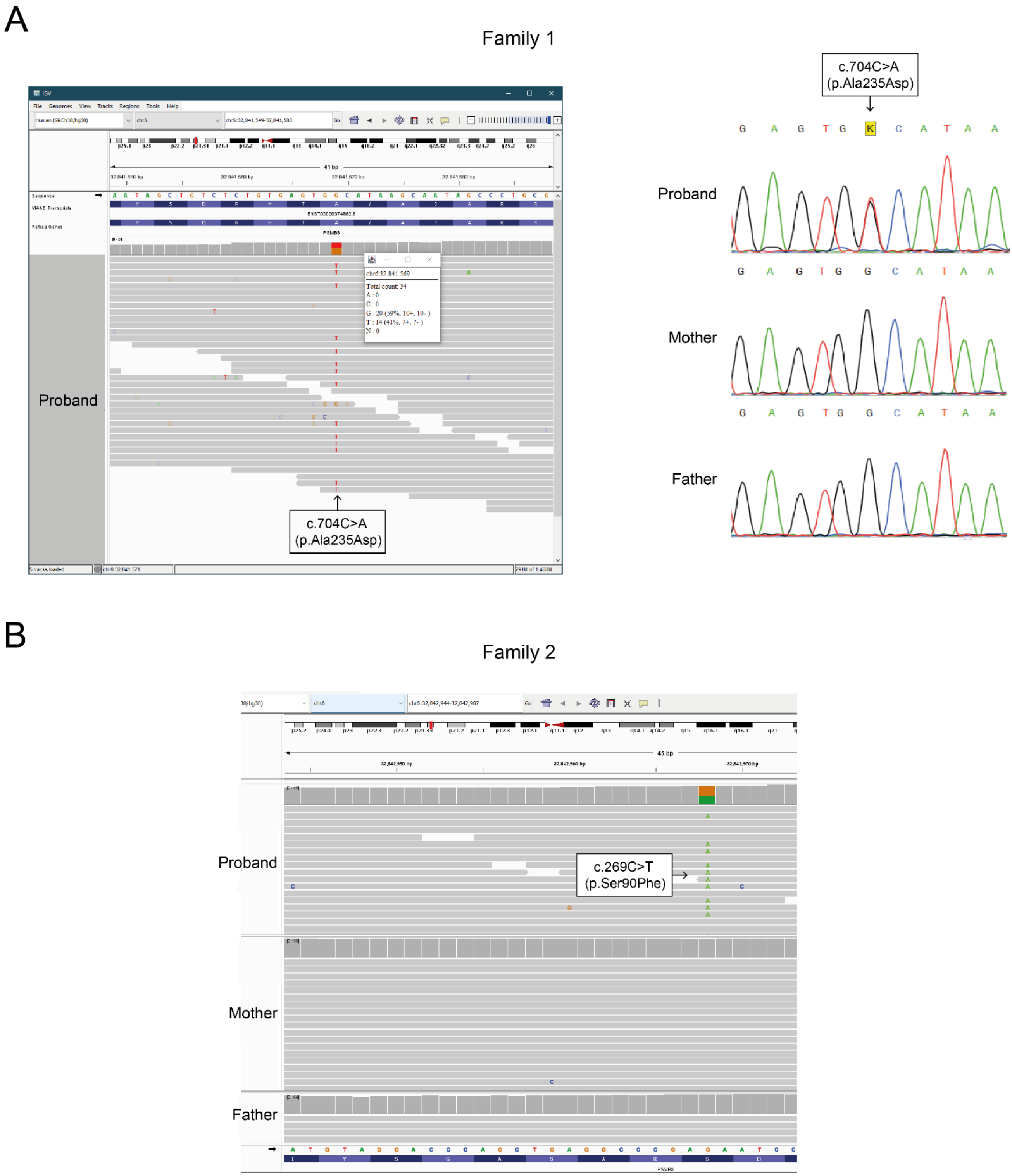

**Supplementary Figure 3. Structural context of p.(Ser90Phe), p.(Ala235Asp), and selected autosomal recessive variants, and associated local frustration changes**

**A**: Variants assessed in this study are highlighted as ball and sticks. Autosomal dominant variants are colored pink while autosomal recessive variants are shown in violet. **B**: Residues that undergo change in frustration energies in the variant p.(Ser90Phe) (neutral/high to minimally frustrated) located 5Å around Ser90 are shown in green. This set also includes the autosomal recessive variant of interest Ala92 shown in purple. Supplementary Table 5 lists these residues. **C**: Residues that undergo change in frustration energies in the variant p.(Ala235Asp) (neutral/minimal to highly frustrated) located 5Å around Ala235 are shown in green. Supplementary Table 5 lists these residues.

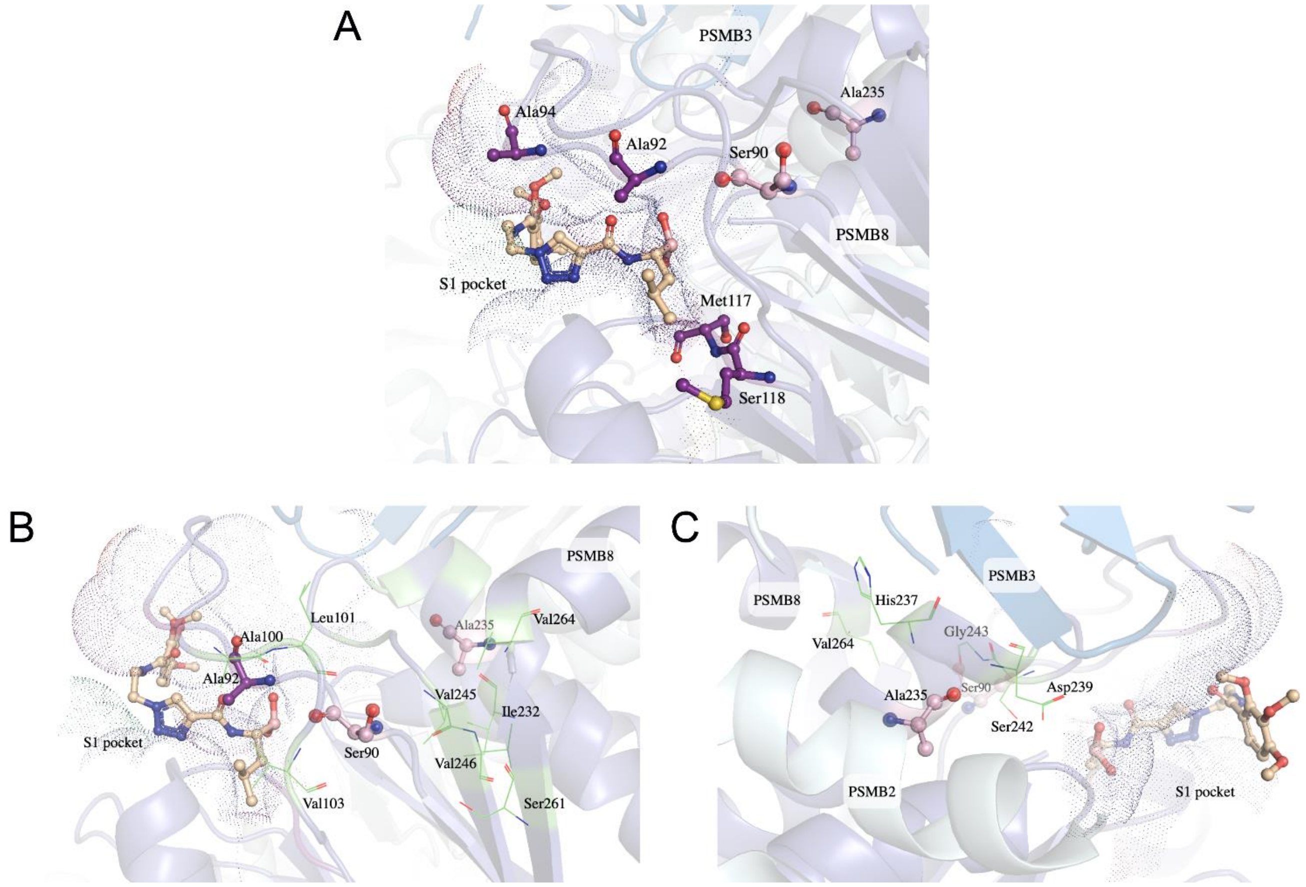

**Supplementary Figure 4. Characterization of standard and immunoproteasome subunits in human osteosarcoma 143B cells and in macrophages derived from THP-1 cells**

**A:** Heatmap representation of the migration profiles of 20S proteasome α- and β-subunits and 19S, 11S (PSME1-2) and PA200 (PSME4) proteasome regulatory proteins reflecting total abundances. Average of three independent experiments. **B:** Quantification of α- and β-subunits. Standard proteasomes predominated in 143B cells whereas in THP-1, immunoproteasomes were more prevalent. iBAQ, intensity-based abundance quantification. **C:** Quantification of the percentage of immunoproteasome content in both cell lines.

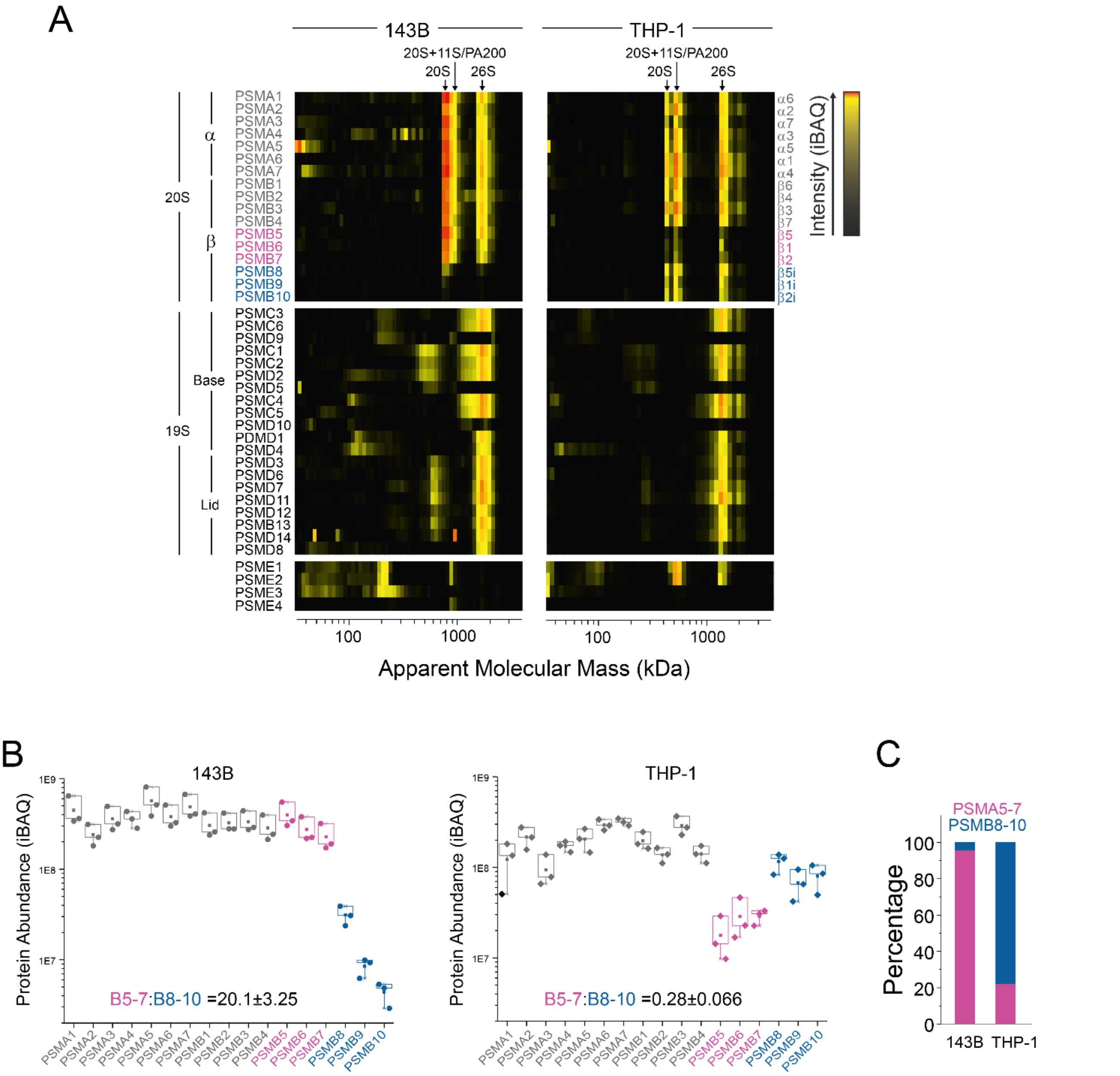

**Supplementary Figure 5. Enhanced expression of immunoproteasome-specific subunits in cytokine-stimulated human skin fibroblasts**

**A**: Heatmaps of the average migration profiles of shared (PSMA1-7, PSMB1-4), standard proteasome-specific (PSMB5-7) and immunoproteasome-specific (PSMB8-10) subunits in control fibroblasts without and after 48 h incubation with IFNγ. Average of two independent experiments. **B:** Quantification of the immunoproteasome content in 20S and 26S inferred from SP-specific subunits:IP-specific subunits abundance ratios in gel fractions with the highest intensity values of 20S and 26S complexes. **C**: Quantification of the percentage of immunoproteasome content after IFNγ stimulation.

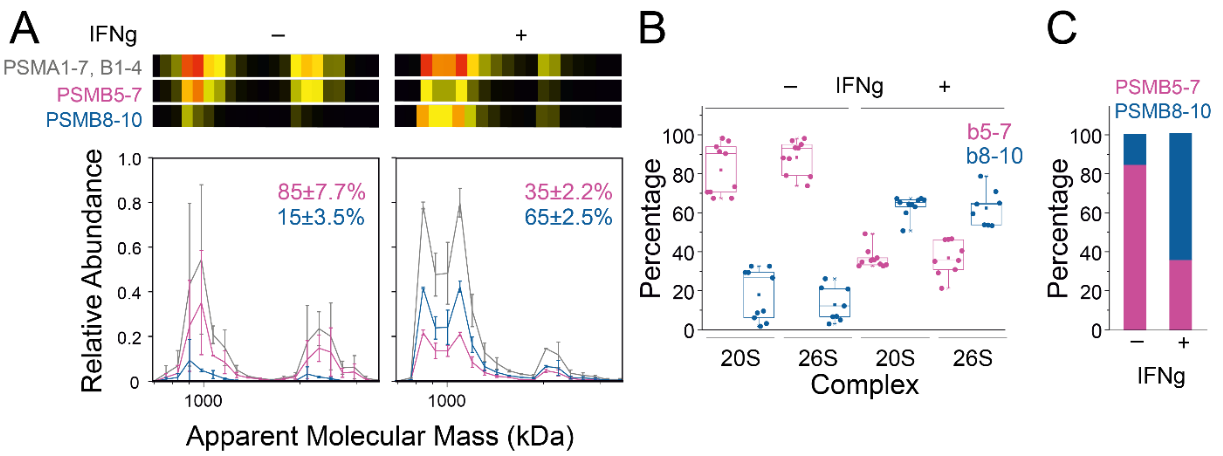

#### Supplementary Figure 6. Heatmaps of the fibroblasts and assembly model

**A:** Heatmap representation of the migration profiles of proteasome subunits and assembly factors in control fibroblast cells and the p.Ala235Asp fibroblast cell line. Average of two independent experiments. **B:** Model of proteasome assembly based on intermediates and fully assembled proteasome complexes evidenced by complexome profiling. Subunits forming assembly intermediates are indicated as cartoon representations, based on cryo-EM structures (PDB: 5GJR (18), 6E5B (19) and 8QYJ (20)). AF: assembly factors.

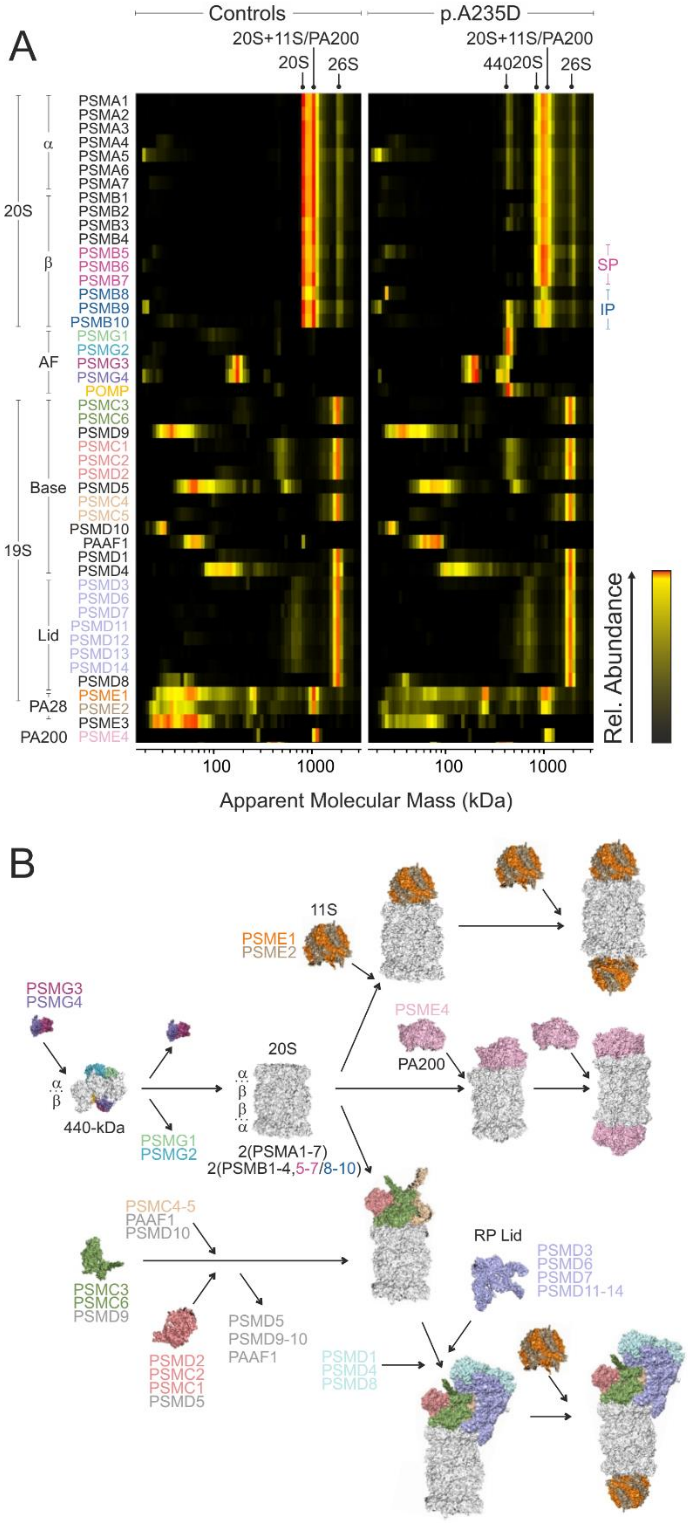

#### Supplementary Figure 7. PSMB8 peptide profiles

**A**: PSMB8 protein sequence with the propeptide sequence highlighted in light pink. Detected peptides corresponding to the propeptide are shown in red, while those from the mature form are shown in black. Identified peptide containing the cleavage site is underscored. **B**: Heatmap representation of the peptides present in the proteomics data in the control and p.(Ala235Asp) samples, with the peptides in the same colors as in A. Average of two independent experiments. **C**: Average migration profiles from precursor-unique peptides and from mature PSMB8 peptides shown in panel B. **D:** Heatmap representation and migration profiles of the wild-type and variant peptides. Average of two independent experiments.

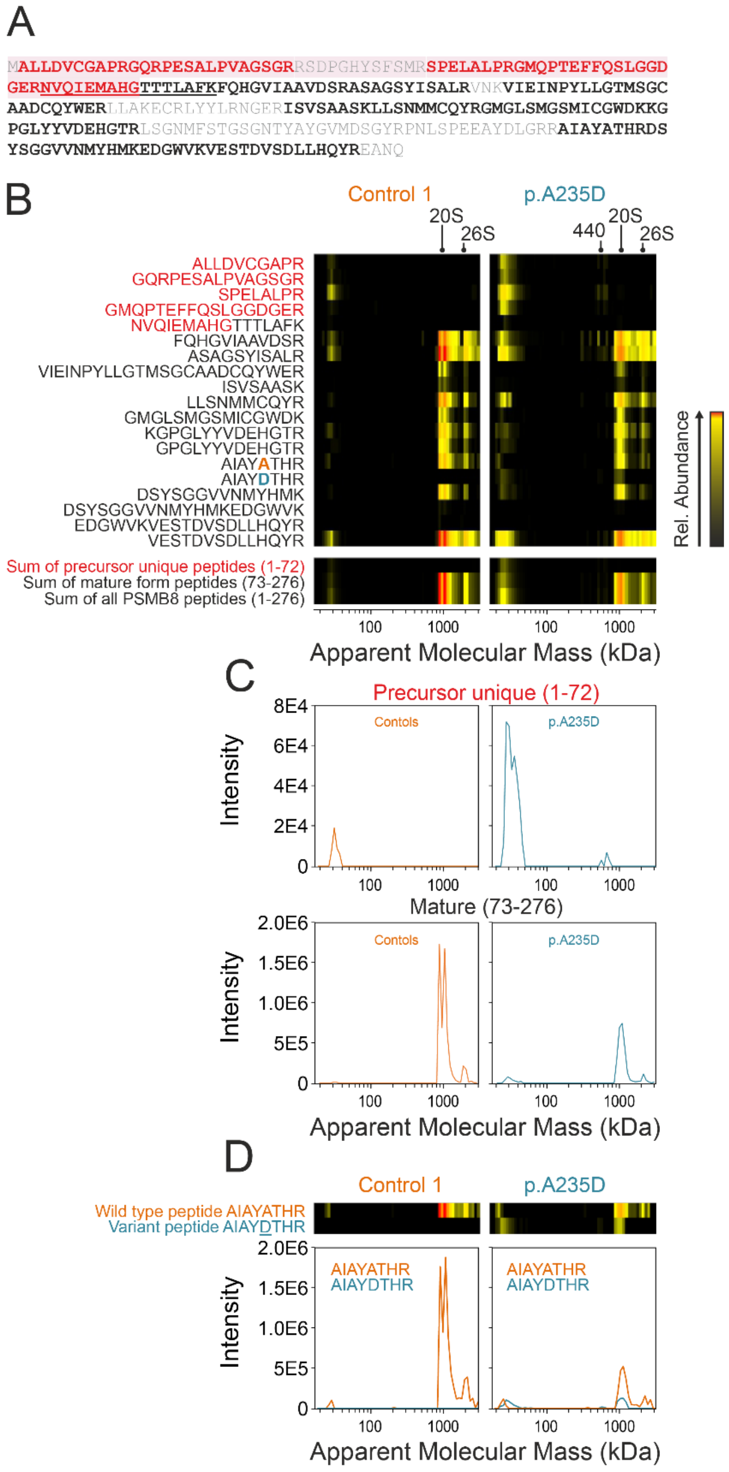

#### Supplementary Figure 8. PSMB9 and PSMB10 peptide profiles

**A**: PSMB9 and PSMB10 protein sequences with the propeptide sequences highlighted in light pink. Detected premature-unique peptides are shown in red, while those from the mature proteins are shown in black. **B**: Heatmap representation of the PSMB9 and PSMB10 peptides present in the proteomics data in the control and p.(Ala235Asp) samples, with the peptides in the same colors as in A. Average of two independent experiments. **C**: Average migration profiles from precursor-unique peptides and mature PSMB9 and PSMB10 peptides.

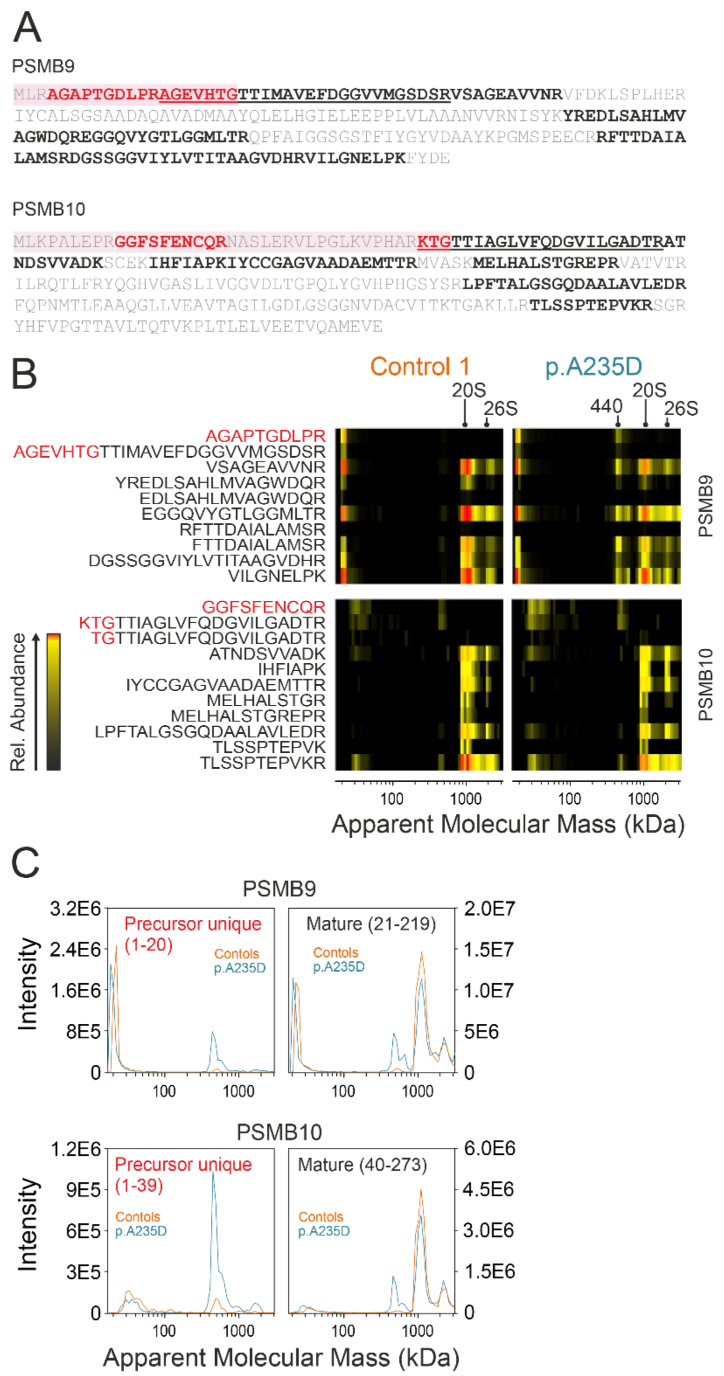

#### Supplementary Figure 9. Gel image following blue native electrophoresis and molecular mass calibration of the fibroblast experiment

**A:** Gel image showing protein separation by blue native electrophoresis and Coomassie staining. **B:** Molecular mass calibration using a reference set of soluble and membrane protein complexes with known molecular masses and stoichiometries. The following proteins and complexes were used to calculate the apparent molecular masses of globular (hydrophilic) protein complexes in each gel fraction: monomeric ATP synthase subunit beta (ATP5F1B, 51.7 kDa), tetrameric single-stranded DNA-binding protein (SSBP1, 15.2×4=61 kDa), dimeric citrate synthase (CS, 49×2=98 kDa), complex I (CI) Q-module (NDUFS2, NDUFS3, NDUFS7, NDUFS8, NDUFA5, NDUFAF3, NDUFAF4, 170 kDa), UDP-glucose:glycoprotein glucosyltransferase 1 (UGGT1, 173 kDa), aldehyde dehydrogenase X tetramer (ALDH1B1, 55.3×4=221 kDa), 60 kDa heat shock protein heptamer (HSPD1, 58×7=406 kDa), isocitrate dehydrogenase [NAD] (IDH3A, IDH3B, IDH3G heterooctamer 300 kDa), propionyl-CoA carboxylase (PCCA, PCCB heterododecamer, 832 kDa), TRiC chaperonin (TCP1, CCT2-5, CCT6A/B, CCT7-8 heterohexadecamer, 944 kDa), alpha ketoglutarate dehydrogenase complex (OGDH, DLST, DLD, MRPS36, ca. 3500 kDa). The following transmembrane protein complexes were used as standards for hydrophobic protein complexes: MIC27 monomer (26 kDa), SLC25A5 (33 kDa), VDAC1-2 dimer (62 kDa), TOMM70 (67 kDa), succinate dehydrogense complex (143 kDa), cytochrome *c* oxidase (CIV, 214 kDa), VDAC1-3 nonamers (290 kDa), cytochrome $bc_1$ complex dimer ($CIII_2$, 485 kDa), ATP synthase (626 kDa), supercomplex $CIII_2$-CIV (700 kDa), supercomplex CI-$CIII_2$ (1485 kDa), respirasome CI-$CIII_2$-CIV (1708 kDa).

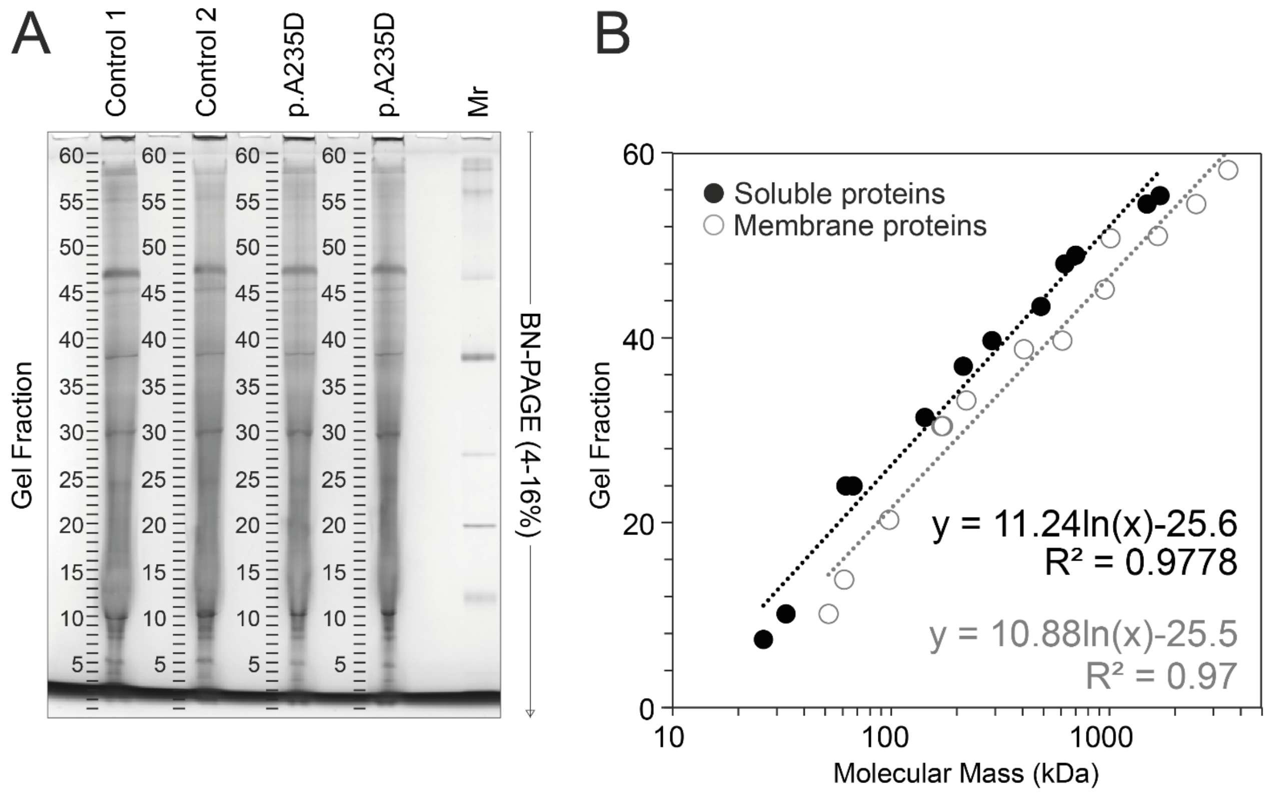

#### Supplementary Tables

##### Supplementary Table 1. Extended laboratory results

| | Individual 1 | | Individual 2 | |
|---|---|---|---|---|
| **Biochemical results** | **Results*** | **Reference value** | **Results*** | **Reference value** |
| C-reactive protein, mg/dL | 91.86 (flare) | 0-5 | 0-8.7 | <1 |
| Ferritin, µg/L | 194-3095 (flare) | 7-140 | 8-112 | 15-300 |
| ALT, U/L | 60 (baseline), 2338 (flare) | <45 | 14-26 | 10-41 |
| AST, U/L | 54 (baseline), 4400 (flare) | <35 | 37-118 | 15-40 |
| CK, U/L | 61-98 (baseline), 219-11.000 (flare) | 34-154 | 21-95 | 55-215 |
| Triglycerides, mg/dL | 187-345 | <100 | 74-147 | 45-203 |
| Cholesterol, mg/dL | 152 | <200 | 188-224 | 112-208 |
| Cholesterol-HDL, mg/dL | 32 | >50 | 38-53 | 38-75 |
| Cholesterol-LDL, mg/dL | 112 | <130 | 120-162 | 64-130 |
| Creatinine, mg/dL | 0.34 | 0.3-1 | 0.29-0.34 | 0.50-0.80 |
| **Blood count** | | | | |
| Red blood cell count, $10^6$/µL | 3.92 | 3.7-4.9 | 3.82-4.26 | 4.03-5.29 |
| Platelet count, $10^3$/µL | 247 | 150-450 | 164-215 | 175-332 |
| White cell blood count, $10^3$/µL | 2.88 | 5.5-15.5 | 2.99-3.04 | 3.84-9.84 |
| Lymphocytes, $10^3$/µL | 1.66 | 2.0-8.0 | 0.91-1.12 | 0.97-3.26 |
| Neutrophils, $10^3$/µL | 0.77 | 1.5-8.5 | 0.58-1.83 | 1.54-7.04 |
| Monocytes, $10^3$/µL | 0.29 | 0-0.8 | 0.39-0.67 | 0.18-0.78 |
| Eosinophils, $10^3$/µL | 0.16 | 0-0.45 | 0.22-0.74 | 0.04-0.38 |
| **Interferon signature** | | | | |
| IFN-signature | 21.64 | <9.4 | 3.64 | <2.82 |
| **General lymphocyte populations** | | | | |
| CD19+, % | 3 | 8.4-28.5 | 6.3 | 13-27 |
| CD19+, /µL | NA | | 131 | 270-860 |
| CD3+, % | 74 | 57.6-81.2 | 90.6 | 60-76 |
| CD3+, /µL | NA | | 1877 | 1200-1600 |
| CD3-CD16/CD56+, % | 21 | 3.5-22.5 | 2.8 | 4-17 |
| CD3-CD16/CD56+, /µL | NA | | 58 | 100-480 |
| CD3+CD4+, % | 49 | 23.6-52.5 | 36.9 | 31-47 |
| CD3+CD4+, /µL | NA | | 765 | 650-1500 |
| CD3+CD8+, % | 21 | 12.1-35.7 | 51.2 | 18-35 |
| CD3+CD8+, /µL | NA | | 1061 | 370-1100 |
| CD4/CD8 ratio | 2.3 | | 0.72 | |
| **B-lymphocyte subpopulations** | | | | |
| CD27-IgM+IgD+, %CD19 | ND | | 84.6 | 43-89 |
| CD27+, %CD19+ | 11 | NA | 11.2 | 2-36 |
| CD27+IgM+IgD+, %CD19+ | 2.75 | 2.9-31.9 | 10.6 | 0-19 |
| CD27+IgM+IgD-, %CD19+ | 2 | NA | 0.1 | 0-5 |
| CD27+IgM-IgD-, %CD19+ | 5.8 | 4.8-20.5 | 0.4 | 0-22 |

| | | | | |
|---|---|---|---|---|
| CD20+, %CD19+ | ND | | 97.1 | 96-100 |
| CD38+Bright IgM+Bright, %CD19+ | ND | | 12 | 0-6 |
| CD38+Bright IgM-, %CD19+ | ND | | 0.4 | 0-3 |
| CD38+Bright, CD19+ | ND | | 17.6 | 0-14 |
| CD21+, %CD19+ | 77 | NA | 97.2 | 86-100 |
| CD21+CD38+, %CD19+ | 53 | NA | 93.1 | 57-96 |
| CD21-CD38+, %CD19+ | 13 | NA | ND | |
| CD21-/low CD38-/low, %CD19+ | ND | | 0.6 | 0-7 |
| **T-lymphocyte subpopulations** | | | | |
| TCR αβ, %CD3+ | 97 | 80-96.7 | ND | |
| TCR γδ, %CD3+ | 3 | 3.4-19.9 | ND | |
| CD4+CD45RA, %CD3+ | 45 | 54.9-83.1 | ND | |
| CD4+CD45RO, %CD3+ | 20 | 12.8-42.5 | ND | |
| CD54RA+CD31+, %CD3+CD4+ | 57 | 47.9-77 | ND | |
| Mitogen response (PHA) | Normal response | | Normal response | |

* Values with "–" indicate ranges based on different determinations. PHA: phytohemagglutinin; NA: not available; ND: not determined.

**Supplementary Table 2. Quantification of soluble serum factors in individual 1 and healthy controls.**

Presence of soluble factors in serum is presented in pg/mL. Serum of healthy controls was used as reference (n=13). Quantification is expressed as mean value (pg/mL) with the lower and upper limit of normal (min-max). Substantial differences of soluble factors in the serum of individual 1 are presented as >1.5x upper limit of normal and highlighted in bold.

| Analyte | Detection range (pg/mL) | Healthy controls (n=13) | | Individual 1 (pg/mL) |
|---|---|---|---|---|
| | | Mean (pg/mL) | Range min-max (pg/mL) | |
| BCA-1 (CXCL13) | 0.64-7236.45 | 12.74 | 5.37-20.80 | **81.58** |
| MIP-4 (CCL18) | 1.58-29290.33 | 5696.71 | 2820.33-13276.04 | **>29290.33** |
| GM-CSF | 1.20-21428.22 | 12.93 | <1.20-59.85 | 2.46 |
| IFNγ | 1.98-43870.11 | 68.93 | 2.20-152.1 | **252.36** |
| IL-1β | 1.88-25072.30 | 25.63 | <1.88-25.63 | <1.88 |
| IL-2 | 0.57-9930.82 | 61.03 | <0.57-61.03 | <0.57 |
| sIL-2Rα | 31.85-499286.07 | 335.83 | 140.86-671.02 | **3256.19** |
| IL-4 | 0.70-10000.07 | 37.35 | 21.16-83.11 | 24.88 |
| IL-5 | 0.64-9960.89 | 4.73 | <0.64-20.10 | 1.52 |
| IL-6 | 0.67-9987.75 | 4.55 | <0.67-8.93 | **20.75** |
| IL-7 | 0.64-10248.88 | 3.75 | <0.64-11.17 | 4.18 |
| IL-8 | 0.70-9297.80 | 5.21 | 1.68-10.19 | **51.19** |
| sVEGFR-2 | 156.33-2526361.67 | 6048.48 | 4762.68-7791.11 | 10012.35 |
| sVCAM-1 | 7.72-124288.47 | 33383.71 | 22807.81-51553.40 | **>124288.47** |
| IL-17A | 3.44-10010.08 | <3.44 | <3.44 | <3.44 |
| IL-21 | 60.79-999925.21 | 3163.99 | <60.79-12873.64 | 181.99 |
| sTNFR-2 | 16.01-216001.48 | 2642.87 | 1488.05-4492.27 | **7244.53** |
| IL-18 | 0.59-10027.34 | 21.90 | 8.94-45.95 | **1214.49** |
| VEGF-D | 45.35-24999.97 | 215.88 | 47.01-342.66 | 82.78 |
| IP-10 (CXCL10) | 1.26-3953.49 | 33.03 | 13.82-63.37 | **752.06** |
| MIP-3 (CCL23) | 0.30-4685.15 | 35.11 | 16.00-80.03 | **>4685.15** |
| MIG (CXCL9) | 1.62-19668.56 | 70.50 | 22.15-150.69 | **897.43** |
| RANTES (CCL5) | 0.53-9706.75 | 1936.65 | 842.65-4136.19 | 932.9 |
| TNFα | 3.21-48196.15 | 15.04 | 5.58-33.21 | 24.45 |
| PF4 (CXCL4) | 1480.00-1057120.00 | 9425.98 | 2388.34-17091.7 | 5278.89 |
| Endostatin | 1.00-4997.00 | 110.83 | 85.12-158.69 | 100.17 |
| TIMP-1 | 238.85-181546.85 | 103681.36 | 76760.81-148601.83 | 90944.74 |

**Supplementary Table 3. Evaluated autoantibodies in individual 1**

| **Autoantibody** | **Individual 1** |
|---|---|
| ANA | Negative |
| ANA: AMA-M2 | Negative |
| ANA: Anti-Histone Antibodies | Negative |
| ANA: Anti-Jo-1 | Negative |
| ANA: Anti-Ribosomal P Protein | Negative |
| ANA: Anti-RNP | Negative |
| ANA: Anti-Scl-70 | Negative |
| ANA: Anti-Smith (Anti-SM) | Negative |
| ANA: Anti-SSA | Negative |
| ANA: Anti-SSB | Negative |
| ANA: DFS70 | Negative |
| ANA: dsDNA | Negative |
| ANA: Anti-Ku | Negative |
| ANA: Anti-Mi-2 | Negative |
| ANA: Anti-Nucleosome | Negative |
| ANA: Anti-PCNA | Negative |
| ANA: Anti-PM-Scl | Negative |
| ANA: Anti-Ro-52 | Negative |
| ANA: Centromere B | Negative |
| ANCA: GBM | Negative |
| ANCA: MPO | Negative |
| ANCA: PR3 | Negative |
| Anti-dsDNA (Crithidia, IFA) | Negative |

#### Supplementary Table 4. Candidate gene variants and rare variants in proteasome subunits

| Individual | Gene | HGVSc | HGVSp | Location | Zygosity | Inheritance | CADD v1.6 | SpliceAI (max) | AF GnomAD v4.0.0 | ClinVar | Comment |
|---|---|---|---|---|---|---|---|---|---|---|---|
| Individual 1 | *PSMB8* | NM_148919.4:c.704C>A | p.(Ala235Asp) | Exon | Heterozygous | De novo | 29.7 | 0.01 | Absent | | |
| | *PSMA5* | NM_002790.4:c.97-897T>C | | Intron | Heterozygous | PV | 15.85 | 0.01 | Absent | | No effect on splicing[a] |
| | *PSMC4* | NM_006503.4:c.674-73G>A | | Intron | Heterozygous | PV | 0.055 | 0.09 | 0.00001001 (0x homozygote) | | No effect on splicing[a] |
| | *CLPB* | NM_030813.6:c.690G>A | p.(Trp230*) | Exon | Heterozygous | PV | 38 | 0.20 | 0.00001115 (0x homozygote) | P 1* | No variant on other allele |
| Individual 2 | *PSMB8* | NM_148919.4:c.269C>T | p.(Ser90Phe) | Exon | Heterozygous | De novo | 28.2 | 0 | Absent | | |
| | *PSMA5* | NM_002790.4:c.97-918A>G | | Intron | Heterozygous | MV | 4.898 | 0 | 0.004326 (5x homozygote) | | No effect on splicing[b] |
| | *PSMA5* | NM_002790.4:c.97-2195G>A | | Intron | Heterozygous | PV | 0.2 | 0 | 0.0004797 (0x homozygote) | | No effect on splicing[b] |
| | *PSMG2* | NM_020232.4:c.289-1235T>C | | Intron | Heterozygous | MV | 2.774 | 0 | 0.00007224 (0x homozygote) | | No effect on splicing[b] |
| | *PSMG2* | NM_020232.4:c.288+2498C>T | | Intron | Heterozygous | MV | 1.376 | 0 | 0.00007223 (0x homozygote) | | No effect on splicing[b] |
| | *PSMB4* | NM_002796.3:c.-1480C>T | | Upstream | Heterozygous | MV | 0.468 | 0 | Absent | | No effect on splicing[b] |
| | *IL25* | NM_022789.3:c.298C>T | p.(Arg100Trp) | Exon | Homozygous | PV MV | 30 | 0.1 | 0.00056322 (1x homozygote) | | Not associated with disease |
| | *GMPPB* | NM_013334.3:c.860G>A | p.(Arg287Gln) | Exon | Heterozygous | PV | 20.9 | 0.02 | 0.00017912 (0x homozygote) | LP/P 2* | No variant on other allele |
| | *HS6ST2* | NM_001077188.1:c.250G>C | p.(Ala84Pro) | Exon | Hemizygous | MV | 21.7 | 0 | Absent | | Does not fit phenotype |

[a]: tested with reverse transcriptase PCR (RT-PCR)

[b]: tested with RNA-seq

LP: likely pathogenic; MV: maternal variant; P: pathogenic; PV: paternal variant

**Supplementary Table 5. Energetic changes and frustration index and for proteasomal contacts across wild-type and mutant variants**

| Variant | Type | 20S (kcal/mol) | 26S (kcal/mol) | Median FI | FI Interpretation | Residue | Chain | TD | ConfFI_HighlyFD | ConfFI_NeutralFD | ConfFI_MinimalFD | MutFI_HighlyFD | MutFI_NeutralFD | MutFI_MinimalFD |
|---|---|---|---|---|---|---|---|---|---|---|---|---|---|---|
| p.Ser90Phe | WT | — | — | -0.671 | Neutral | 18 | K | 44 | 10 | 31 | 3 | 10 | 27 | 7 |
| | | | | | | | Y | 44 | 9 | 32 | 3 | 11 | 27 | 6 |
| | MUT | 13.05 | 6.13 | 1.356 | Minimally frustrated | 18 | K | 44 | 9 | 23 | 12 | 7 | 22 | 15 |
| | | | | | | | Y | 44 | 7 | 25 | 12 | 7 | 22 | 15 |
| p.Ala235Asp | WT | — | — | 0.607 | Neutral | 163 | K | 56 | 6 | 40 | 10 | 0 | 36 | 20 |
| | | | | | | | Y | 56 | 6 | 40 | 10 | 0 | 36 | 20 |
| | MUT | 28.26 | 16.91 | -1.165 | Highly frustrated | 163 | K | 56 | 7 | 41 | 8 | 10 | 33 | 13 |
| | | | | | | | Y | 56 | 7 | 41 | 8 | 10 | 33 | 13 |
| p.Ala92Thr | WT | — | — | 0.734 | Neutral | 20 | K | 31 | 0 | 26 | 5 | 1 | 22 | 8 |
| | | | | | | | Y | 31 | 0 | 26 | 5 | 2 | 21 | 8 |
| | MUT | 1.21 | -0.42 | -0.249 | Neutral | 20 | K | 31 | 0 | 27 | 4 | 1 | 26 | 4 |
| | | | | | | | Y | 31 | 0 | 27 | 4 | 1 | 26 | 4 |
| p.Ala92Val | WT | — | — | 0.734 | Neutral | 20 | K | 31 | 0 | 26 | 5 | 1 | 22 | 8 |
| | | | | | | | Y | 31 | 0 | 26 | 5 | 2 | 21 | 8 |
| | MUT | -0.67 | -2.58 | 1.455 | Minimally frustrated | 20 | K | 31 | 0 | 24 | 7 | 0 | 21 | 10 |
| | | | | | | | Y | 31 | 0 | 24 | 7 | 1 | 20 | 10 |
| p.Ala94Pro | WT | — | — | -0.434 | Neutral | 22 | K | 13 | 0 | 11 | 2 | 0 | 13 | 0 |
| | | | | | | | Y | 13 | 0 | 11 | 2 | 1 | 12 | 0 |
| | MUT | 1.25 | 7.32 | -0.237 | Neutral | 22 | K | 13 | 0 | 10 | 3 | 2 | 9 | 2 |
| | | | | | | | Y | 13 | 0 | 10 | 3 | 1 | 10 | 2 |
| p.Met117Val | WT | — | — | 0.708 | Neutral | 45 | K | 51 | 6 | 38 | 7 | 3 | 31 | 17 |
| | | | | | | | Y | 51 | 7 | 37 | 7 | 4 | 31 | 16 |
| | MUT | 4.39 | 0.37 | 1.14 | Minimally frustrated | 45 | K | 51 | 6 | 36 | 9 | 2 | 31 | 18 |
| | | | | | | | Y | 51 | 7 | 35 | 9 | 3 | 30 | 18 |

| Variant | Type | 20S (kcal/mol) | 26S (kcal/mol) | Median FI | FI Interpretation | Residue | Chain | TD | ConfFI_HighlyFD | ConfFI_NeutralFD | ConfFI_MinimalFD | MutFI_HighlyFD | MutFI_NeutralFD | MutFI_MinimalFD |
|---|---|---|---|---|---|---|---|---|---|---|---|---|---|---|
| p.Ser118Pro | WT | — | — | -0.144 | Neutral | 46 | K | 48 | 0 | 42 | 6 | 2 | 32 | 14 |
| | | | | | | | Y | 48 | 0 | 42 | 6 | 2 | 33 | 13 |
| | MUT | 12.25 | — | -0.154 | Neutral | 46 | K | 49 | 0 | 43 | 6 | 4 | 32 | 13 |
| | | | | | | | Y | 49 | 0 | 43 | 6 | 7 | 32 | 10 |

**Legend**:

- 20S/26S kcal/mol: ΔΔG values estimated using FoldX on the 20S immunoproteasome (PDB: 6E5B) and 26S proteasome structure (PDB: 6MSB), respectively.
- Frustration analysis was performed on the 20S immunoproteasome (PDB: 6E5B), and values are based on calculations from chains K and Y.
- Median FI: Median single-residue frustration index (FI), averaged across chains K and Y.
- ConfFI/MutFI: Local contact densities based on configurational or mutational frustration indices, respectively. Calculated within a 5 Å radius centered at the Cα atom of the residue of interest.
- Residue: Residue number as defined in the PDB structure.
- TD (Total Density): Total number of residue–residue contacts within a 5 Å radius of the Cα atom.
- HighlyFD/NeutralFD/MinimalFD: Fraction of local contacts that are classified as highly, neutrally, or minimally frustrated, respectively, within the 5 Å radius.

**Additional data:**

- For p.Ser90Phe, the MinimalFD values (in both ConfFI and MutFI) represent a total of 16 contacts with 9 residues: Ala92, Ala100, Leu101, Val103, Ile232, Val245, Val246, Ser261, Val264.
- For p.Ala235Asp, the HighlyFD values (in both ConfFI and MutFI) represent 10 contacts with 5 residues: His237, Asp239, Ser242, Gly243, Val264.

**Supplementary Table 6. Enriched GO terms**

The table lists GO terms significantly enriched among the analyzed genes, including the term ID and description, GO category (Biological Process [BP], Cellular Component [CC], or Molecular Function [MF]), the count and percentage of genes in the input list annotated with the term, p-value and false discovery rate (FDR) for enrichment significance, total genes in the input list, total population hits (Pop Hits) and population size (Pop Total) used for the analysis, fold enrichment, and the genes associated with each GO term.

| **Term** | **Category** | **Count** | **%** | **PValue** | **List Total** | **Pop Hits** | **Pop Total** | **Fold Enrichment** | **FDR** | **Genes** |
|---|---|---|---|---|---|---|---|---|---|---|
| GO:0006955~immune response | BP_DIRECT | 14 | 6.542 | 1.12077E-05 | 207 | 98 | 6496 | 4.483 | 0.014760492 | *CXCL9, HLA-DRB4, SEMA3C, IFI44, PIK3R2, TNFAIP1, PDCD1LG2, CRIP1, IFI44L, ACKR3, JAK2, HLA-DRB3, HLA-DQB1, NECTIN1* |
| GO:0007155~cell adhesion | BP_DIRECT | 20 | 9.346 | 3.49961E-05 | 207 | 211 | 6496 | 2.975 | 0.023044909 | *CGREF1, COL15A1, TNXB, MCAM, PCDHGC4, CTNND2, INPPL1, PRKCA, ATP1B1, COMP, ISLR, CEACAM1, S1PR1, SSPN, ACKR3, JAK2, MFGE8, CD34, CTNNAL1, NECTIN1* |
| GO:0016324~apical plasma membrane | CC_DIRECT | 17 | 7.944 | 5.4856E-05 | 212 | 165 | 6719 | 3.265 | 0.015085402 | *OXTR, PATJ, JAG1, MTCL1, ABCB4, FZD6, SLC2A1, ATP1B1, SLC2A5, PLD1, CEACAM1, SLCO3A1, SLC6A9, SLC39A8, STX3, SLC16A3, CD34* |
| GO:0004930~G protein-coupled receptor activity | MF_DIRECT | 7 | 3.271 | 9.43101E-05 | 209 | 25 | 6643 | 8.900 | 0.045551802 | *FZD7, FZD6, F2R, S1PR1, LPAR1, ACKR3, LGR4* |

#### Members of the Undiagnosed Diseases Network (Version 3.31.25)

Alyssa A. Tran, Arjun Tarakad, Ashok Balasubramanyam, Brendan H. Lee, Carlos A. Bacino, Daryl A. Scott, Elaine Seto, Gary D. Clark, Hongzheng Dai, Hsiao-Tuan Chao, Ivan Chinn, James P. Orengo, Jennifer E. Posey, Jill A. Rosenfeld, Kim Worley, Lindsay C. Burrage, Lisa T. Emrick, Lorraine Potocki, Monika Weisz Hubshman, Richard A. Lewis, Ronit Marom, Seema R. Lalani, Shamika Ketkar, Tiphanie P. Vogel, William J. Craigen, Jared Sninsky, Lauren Blieden, Sandesh Nagamani, Hugo J. Bellen, Michael F. Wangler, Oguz Kanca, Shinya Yamamoto, Christine M. Eng, Patricia A. Ward, Pengfei Liu, Adeline Vanderver, Cara Skraban, Edward Behrens, Gonench Kilich, Kathleen Sullivan, Kelly Hassey, Ramakrishnan Rajagopalan, Rebecca Ganetzky, Vishnu Cuddapah, Anna Raper, Daniel J. Rader, Giorgio Sirugo, Vaidehi Jobanputra, Allyn McConkie-Rosell, Kelly Schoch, Mohamad Mikati, Nicole M. Walley, Rebecca C. Spillmann, Vandana Shashi, Alan H. Beggs, Calum A. MacRae, David A. Sweetser, Deepak A. Rao, Edwin K. Silverman, Elizabeth L. Fieg, Frances High, Gerard T. Berry, Ingrid A. Holm, J. Carl Pallais, Joan M. Stoler, Joseph Loscalzo, Lance H. Rodan, Laurel A. Cobban, Lauren C. Briere, Matthew Coggins, Melissa Walker, Richard L. Maas, Susan Korrick, Jessica Douglas, Cecilia Esteves, Emily Glanton, Isaac S. Kohane, Kimberly LeBlanc, Rachel Mahoney, Shamil R. Sunyaev, Shilpa N. Kobren, Brett H. Graham, Erin Conboy, Francesco Vetrini, Kayla M. Treat, Khurram Liaqat, Lili Mantcheva, Stephanie M. Ware, Breanna Mitchell, Brendan C. Lanpher, Devin Oglesbee, Eric Klee, Filippo Pinto e Vairo, Ian R. Lanza, Kahlen Darr, Lindsay Mulvihill, Lisa Schimmenti, Queenie Tan, Surendra Dasari, Abdul Elkadri, Brett Bordini, Donald Basel, James Verbsky, Julie McCarrier, Michael Muriello, Michael Zimmermann, Adriana Rebelo, Carson A. Smith, Deborah Barbouth, Guney Bademci, Joanna M. Gonzalez, Kumarie Latchman, LéShon Peart, Mustafa Tekin, Nicholas Borja, Stephan Zuchner, Stephanie Bivona, Willa Thorson, Herman Taylor, Rakale C. Quarells, Ayuko Iverson, Bruce Gelb, Charlotte Cunningham-Rundles, Eric Gayle, Joanna Jen, Louise Bier, Mafalda Barbosa, Manisha Balwani, Mariya Shadrina, Rachel Evard, Saskia Shuman, Susan Shin, Andrea Gropman, Barbara N. Pusey Swerdzewski, Camilo Toro, Colleen E. Wahl, Donna Novacic, Ellen F. Macnamara, John J. Mulvihill, Maria T. Acosta, Precilla D'Souza, Valerie V. Maduro, Ben Afzali, Ben Solomon, Cynthia J. Tifft, David R. Adams, Elizabeth A. Burke, Francis Rossignol, Heidi Wood, Jiayu Fu, Joie Davis, Leoyklang Petcharet, Lynne A. Wolfe, Margaret Delgado, Marie Morimoto, Marla Sabaii, MayChristine V. Malicdan, Neil Hanchard, Orpa Jean-Marie, Wendy Introne, William A. Gahl, Yan Huang, Andrew Stergachis, Danny Miller, Elisabeth Rosenthal, Elizabeth Blue, Elsa Balton, Emily Shelkowitz, Eric Allenspach, Fuki M. Hisama, Gail P. Jarvik, Ghayda Mirzaa, Ian Glass, Kathleen A. Leppig, Katrina Dipple, Mark Wener, Martha Horike-Pyne, Michael Bamshad, Peter Byers, Runjun Kumar, Seth Perlman, Sirisak Chanprasert, Virginia Sybert, Wendy Raskind, Nitsuh K. Dargie, Chun-Hung Chan, Dr. Francisco Bustos velasq, Isum Ward, Jason Schend, Jennifer Morgan, Megan Bell, Miranda Leitheiser, Mohamad Saifeddine, Paul Berger, Rachel Li, Taylor Beagle, Alexander Miller, Beatriz Anguiano, Beth A. Martin, Brianna Tucker, Chloe M. Reuter, Devon Bonner, Elijah Kravets, Hector Rodrigo Mendez, Holly K. Tabor, Jacinda B. Sampson, Jason Hom, Jennefer N. Kohler, Jennifer Schymick, John E. Gorzynski, Jonathan A. Bernstein, Kevin S. Smith, Laura Keehan, Laurens Wiel, Matthew T. Wheeler, Meghan C. Halley, Mia Levanto, Page C. Goddard, Paul G. Fisher, Rachel A. Ungar, Raquel L. Alvarez, Sara Emami, Shruti Marwaha, Stephen B Montgomery, Suha Bachir, Tanner D Jensen, Taylor Maurer, Terra R. Coakley, Euan A. Ashley, Ali Al-Beshri, Anna Hurst, Brandon M Wilk, Bruce Korf, Elizabeth A Worthey, Kaitlin Callaway, Martin Rodriguez, Tammi Skelton, Tarun KK Mamidi, Andrew B. Crouse, Jordan Whitlock, Mariko Nakano-Okuno, Matthew Might, William E. Byrd, Albert R. La Spada, Changrui Xiao, Elizabeth C. Chao, Eric Vilain, Jose Abdenur, Kirsten Blanco, Maija-Rikka Steenari, Rebekah Barrick, Richard Chang, Sanaz Attaripour, Suzanne Sandmeyer, Tahseen Mozaffar, Alden Huang, Andres Vargas, Bianca E. Russell, Brent L. Fogel, Esteban C. Dell'Angelica, George Carvalho, Julian A. Martínez-Agosto, Layal F. Abi Farraj, Manish J. Butte, Martin G. Martin, Naghmeh Dorrani, Neil H. Parker, Rosario I. Corona, Stanley F. Nelson, Yigit Karasozen, Aaron Quinlan, Alistair Ward, Ashley Andrews, Corrine K. Welt, Dave Viskochil, Erin E. Baldwin, John Carey, Justin Alvey, Laura Pace, Lorenzo Botto, Nicola Longo, Paolo Moretti, Rebecca Overbury, Russell Butterfield, Steven Boyden, Thomas J. Nicholas, Matt Velinder, Gabor Marth, Pinar Bayrak-Toydemir, Rong Mao, Monte Westerfield, Brian Corner, John A. Phillips III, Kimberly Ezell, Lynette Rives, Rizwan Hamid, Serena Neumann, Ashley McMinn, Joy D. Cogan, Thomas Cassini, Alex Paul, Dana Kiley, Daniel Wegner, Erin McRoy, Jennifer Wambach, Kathy Sisco, Patricia Dickson, F. Sessions Cole, Dustin Baldridge, Jimann Shin, Lilianna Solnica-Krezel, Stephen C. Pak, Timothy Schedl, Allen Bale, Carol Oladele, Caroline Hendry, Emily Wang, Hua Xu, Hui Zhang, Lauren Jeffries, María José Ortuño Romero, Mark Gerstein, Michele Spencer-Manzon, Monkol Lek, Nada Derar, Odelya Kaufman, Shrikant Mane, Teodoro Jerves Serrano, Vasilis Vasiliou, Winston Halstead, Yong-Hui Jiang

#### References

1. Vasimuddin M, Misra S, Li H, Aluru S. Efficient Architecture-Aware Acceleration of BWA-MEM for Multicore Systems. Int Parall Distrib P. 2019:314-24.
2. Okonechnikov K, Conesa A, Garcia-Alcalde F. Qualimap 2: advanced multi-sample quality control for high-throughput sequencing data. Bioinformatics. 2016;32(2):292-4.
3. McKenna A, Hanna M, Banks E, Sivachenko A, Cibulskis K, Kernytsky A, et al. The Genome Analysis Toolkit: A MapReduce framework for analyzing next-generation DNA sequencing data. Genome Research. 2010;20(9):1297-303.
4. Chen X, Schulz-Trieglaff O, Shaw R, Barnes B, Schlesinger F, Kallberg M, et al. Manta: rapid detection of structural variants and indels for germline and cancer sequencing applications. Bioinformatics. 2016;32(8):1220-2.
5. Roller E, Ivakhno S, Lee S, Royce T, Tanner S. Canvas: versatile and scalable detection of copy number variants. Bioinformatics. 2016;32(15):2375-7.
6. Dolzhenko E, van Vugt J, Shaw RJ, Bekritsky MA, van Blitterswijk M, Narzisi G, et al. Detection of long repeat expansions from PCR-free whole-genome sequence data. Genome Res. 2017;27(11):1895-903.
7. Huijser E, Bodewes ILA, Lourens MS, van Helden-Meeuwsen CG, van den Bosch TPP, Grashof DGB, et al. Hyperresponsive cytosolic DNA-sensing pathway in monocytes from primary Sjogren's syndrome. Rheumatology (Oxford). 2022;61(8):3491-6.
8. Bodewes ILA, Al-Ali S, van Helden-Meeuwsen CG, Maria NI, Tarn J, Lendrem DW, et al. Systemic interferon type I and type II signatures in primary Sjogren's syndrome reveal differences in biological disease activity. Rheumatology (Oxford). 2018;57(5):921-30.
9. Rice GI, Forte GM, Szynkiewicz M, Chase DS, Aeby A, Abdel-Hamid MS, et al. Assessment of interferon-related biomarkers in Aicardi-Goutieres syndrome associated with mutations in TREX1, RNASEH2A, RNASEH2B, RNASEH2C, SAMHD1, and ADAR: a case-control study. Lancet Neurol. 2013;12(12):1159-69.
10. Wittig I, Braun HP, Schägger H. Blue native PAGE. Nat Protoc. 2006;1(1):418-28.
11. Perez-Riverol Y, Bandla C, Kundu DJ, Kamatchinathan S, Bai JW, Hewapathirana S, et al. The PRIDE database at 20 years: 2025 update. Nucleic Acids Research. 2024;53(D1):D543-D53.
12. Demichev V, Messner CB, Vernardis SI, Lilley KS, Ralser M. DIA-NN: neural networks and interference correction enable deep proteome coverage in high throughput. Nat Methods. 2020;17(1):41-4.
13. de Hoon MJ, Imoto S, Nolan J, Miyano S. Open source clustering software. Bioinformatics. 2004;20(9):1453-4.
14. Giese KP, Aziz W, Kraev I, Stewart MG. Generation of multi-innervated dendritic spines as a novel mechanism of long-term memory formation. Neurobiol Learn Mem. 2015;124:48-51.
15. Van Strien J, Guerrero-Castillo S, Chatzispyrou IA, Houtkooper RH, Brandt U, Huynen MA. COmplexome Profiling ALignment (COPAL) reveals remodeling of mitochondrial protein complexes in Barth syndrome. Bioinformatics. 2019;35(17):3083-91.
16. van Strien J, Haupt A, Schulte U, Braun HP, Cabrera-Orefice A, Choudhary JS, et al. CEDAR, an online resource for the reporting and exploration of complexome profiling data. Biochim Biophys Acta Bioenerg. 2021;1862(7):148411.
17. Dong Y, Zhang S, Wu Z, Li X, Wang WL, Zhu Y, et al. Cryo-EM structures and dynamics of substrate-engaged human 26S proteasome. Nature. 2019;565(7737):49-55.
18. Huang X, Luan B, Wu J, Shi Y. An atomic structure of the human 26S proteasome. Nat Struct Mol Biol. 2016;23(9):778-85.
19. Ladi E, Everett C, Stivala CE, Daniels BE, Durk MR, Harris SF, et al. Design and Evaluation of Highly Selective Human Immunoproteasome Inhibitors Reveal a Compensatory Process That Preserves Immune Cell Viability. J Med Chem. 2019;62(15):7032-41.
20. Adolf F, Du J, Goodall EA, Walsh RM, Jr., Rawson S, von Gronau S, et al. Visualizing chaperone-mediated multistep assembly of the human 20S proteasome. Nat Struct Mol Biol. 2024;31(8):1176-88.
